# Large artery phenotypes, cerebrovascular function, and progression of cerebral small vessel disease

**DOI:** 10.64898/2026.05.23.26344314

**Authors:** Fei Han, Una Clancy, Carmen Arteaga-Reyes, Michael J. Thrippleton, Maria Del C. Valdés Hernández, Daniela Jaime Garcia, Michael S. Stringer, Ellen Backhouse, Francesca M. Chappell, Yajun Cheng, Dillys Xiaodi Liu, Junfang Zhang, Angela C.C. Jochems, Eleni Sakka, Charlotte Jardine, Gayle Barclay, Donna McIntyre, Iona Hamilton, Rosalind Brown, Fergus N. Doubal, Yi-Cheng Zhu, Joanna M. Wardlaw

## Abstract

**Objective:** Cranial artery stenosis and dilatation are distinct large artery phenotypes that often coexist with cerebral small vessel disease (cSVD), yet their downstream microvascular functional correlates remain unclear.

**Methods:** In the prospective Mild Stroke Study 3, we recruited patients with lacunar or mild non-lacunar stroke. At baseline, large artery stenosis (LAS), basilar artery dolichoectasia (BADE), and intracranial arterial diameters were assessed. Multimodal MRI quantified cerebrovascular reactivity (CVR), blood-brain barrier (BBB) permeability, plasma volume fraction, and intracranial pulsatility. cSVD markers were evaluated at baseline and 1 year. Associations between large artery phenotypes and vascular function were examined with multivariable regression. Mediation analyses tested whether vascular dysfunction linked large artery pathology to cSVD progression.

**Results:** Among 224 participants (mean age 66.0 ± 11.2 years; 66.5% men), BADE (*n*=36, 16.1%) was independently associated with lower CVR in normal-appearing white matter (NAWM; β −0.01, 95% CI −0.016 to −0.004, *P*=0.003). Larger mean intracranial arterial diameter was associated with lower CVR in NAWM and white matter hyperintensities (WMH), while showing a U-shaped association with BBB permeability. LAS (*n*=46, 20.5%) was unrelated to CVR, BBB permeability, or pulsatility, but was associated with higher plasma volume in WMH. CVR in NAWM partially mediated the association between BADE and both baseline cSVD burden and 1-year progression.

**Interpretation:** Large artery dilatation may serve as a macroscopic signal of small-vessel dysfunction, being associated with lower CVR and altered BBB permeability. Reduced CVR in NAWM partially mediated the impact of dolichoectasia on cSVD progression and may represent a potential therapeutic target.

---

The cerebral circulation constitutes a continuous vascular tree in which alterations in large arteries and the microvasculature frequently co-exist, reflecting exposure to shared vascular risk factors and genetic susceptibility. Large artery stenosis (LAS) may reduce distal perfusion pressure and disturb flow dynamics, whereas intracranial dolichoectasia, characterized by arterial dilatation and tortuosity, is regarded as a manifestation of vascular wall changes with loss of elasticity and impaired compliance.^1, 2^ In our and other’s previous work, we demonstrated that these large artery phenotypes have divergent aetiopathological implications for stroke subtype and cerebral small vessel disease (cSVD): LAS was uncommon in lacunar stroke and unrelated to cSVD markers, while dilatation related strongly to lacunar stroke and cSVD.^3^

cSVD accounts for up to one quarter of ischemic strokes and is a leading cause of vascular cognitive impairment.^4–6^ Patients with cSVD consistently exhibit vascular dysfunction, including reduced cerebrovascular reactivity (CVR),^7–9^ increased blood-brain barrier (BBB) permeability,^10, 11^ and increased intracranial vascular pulsatility.^12–14^ Advances in multimodal MRI enable these processes to be quantified in vivo. Yet it remains unclear whether, and to what extent, upstream large-artery phenotype is linked with these tissue-level vascular functional disturbances. Clarifying this relationship is critical to understanding how conduit artery pathology may signal microvascular failure and subsequent tissue injury.

To address this gap, we conducted a multimodal MRI study within the Mild Stroke Study 3 (MSS3), a prospective cohort of lacunar stroke, with mild non-lacunar stroke as a comparator, enriched for cSVD features. Using reproducible validated imaging techniques, we quantified CVR, BBB permeability, and pulsatility, and examined their associations with cranial artery stenosis on one hand, and with arterial dilatation, characterized by basilar artery dolichoectasia (BADE) and intracranial arterial diameters, on the other. We further investigated whether impaired vascular function mediated the relationship between large artery pathology and cSVD-related structural brain features, including baseline burden and lesion progression over 1 year.

## Methods

This study follows the Strengthening the Reporting of Observational Studies in Epidemiology (STROBE) guidelines.^15^

### Participants and study design

We analyzed data from the MSS3, a prospective cohort of patients with mild ischemic stroke. The study protocol is published.^16^ In brief, we consecutively recruited patients presenting to Edinburgh stroke services between 2018 to 2021 with functionally independent ischemic stroke (National Institutes of Health Stroke Scale <8 and modified Rankin scale ≤2 at recruitment), including both lacunar and non-lacunar (minor cortical) subtypes. Stroke diagnosis and subtyping were conducted by stroke specialists. Patients were followed for 1 year, up to 2022. All patients received secondary prevention therapies according to UK stroke guidelines, including antiplatelet, lipid-lowering, and antihypertensive drugs as appropriate. Non-lacunar strokes served as the comparator group because they shared similar risk factor profiles and received standardized treatment, thereby minimizing potential confounding effects of medication on vascular function measures and enabling investigation of cSVD associations across a broad spectrum of severities. We excluded individuals with contraindications to MRI or with severe neurological, cardiac, or respiratory comorbidities. Comprehensive clinical and neuroimaging assessments were performed at baseline and repeated at 6- and 12- month follow-up visits (Figure 1).

**Figure 1.**
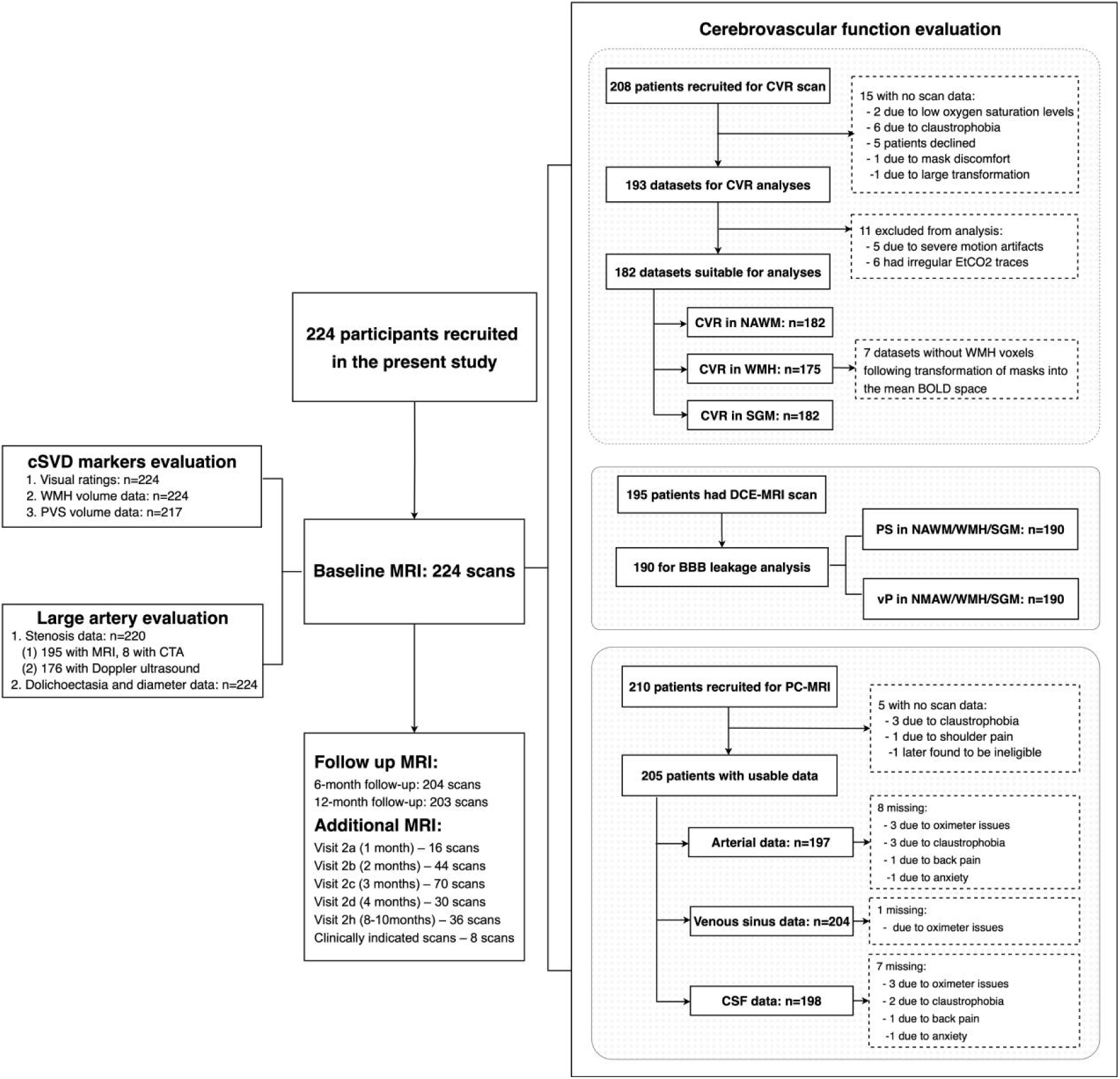
Flowchart of the study. Abbreviations: BOLD, blood oxygen level-dependent; DCE, dynamic contrast-enhanced magnetic resonance imaging; MRI, magnetic resonance image; CTA: computed tomography angiography; CVR, cerebrovascular reactivity; PS, permeability surface area product; *v_p_*, plasma volume; NAWM, normal-appearing white matter; WMH, white matter hyperintensity; SGM, subcortical gray matter.

### Standard Protocol Approvals, Registrations, and Patient Consents

This study was approved by the Southeast Scotland Regional Ethics Committee (reference.18/SS/0044) and adhered to the Declaration of Helsinki. Written informed consent was obtained from all participants. The study is registered under ISRCTN 12113543.

### MRI acquisition

All participants underwent diagnostic imaging at the initial stroke presentation. During baseline visit (1-3 months post-stroke, to minimize acute effects), structural and vascular function scans were performed on the same 3T MRI scanner (Siemens Prisma; Siemens Healthcare, Erlangen, Germany). Full details of the MRI acquisition protocols are published.^16, 17^ In brief, the protocol included: (1) Structural imaging: 3D T1-weighted (T1W), T2-weighted (T2W), fluid attenuated inversion recovery (FLAIR), susceptibility-weighted imaging (SWI), and 2D diffusion-weighted imaging (DWI) to assess cSVD burden and brain volumes; (2) Dynamic blood oxygen level-dependent (BOLD) sequence: acquired during alternating inhalation of 2 minutes medical air and 3 minutes 6% CO_2_, a validated paradigm for assessing CVR in cSVD that elicits robust microvascular responses under natural, unforced respiration; (3) Dynamic contrast-enhanced (DCE) MRI: performed following intravenous gadobutrol (0.1 mmol/kg; 1M Gadovist, Bayer AG, Leverkusen, Germany) to assess BBB leakage (permeability surface area product [PS]) and plasma volume fraction (*v_p_*); (4) Phase contrast (PC) MRI: to measure blood flow and pulsatility in the internal carotid and vertebral arteries, internal jugular veins, straight, sagittal and transverse venous sinuses, and cerebrospinal fluid (CSF) at the foramen magnum.

All follow-up MRI examinations were performed using the same scanner and sequence parameters as baseline, under a standardized quality assurance program. Vascular function measurements were collected at baseline only.

### MRI processing and analysis

All image processing and analyses followed standardized protocols and were conducted by an experienced team under the supervision of a neuroradiologist (J.M.W.). Investigators were blinded to clinical information. Large artery assessments, structural MRI analysis, and vascular function processing were performed independently by different researchers at separate time points, each blinded to the other measures. Detailed processing methods are provided in the Supplementary Methods or the published protocol.^3, 16^

#### Large artery assessment

LAS was defined as ≥50% stenosis in any intracranial large artery or the cervical internal carotid artery (ICA), assessed using DCE-MRI, carotid ultrasound, and available angiographic imaging according to established criteria (Supplementary Methods and Figure S1).^18, 19^ Intracranial artery diameters were measured on axial T2W images for seven arteries: bilateral ICAs (cavernous segment), bilateral middle cerebral arteries (MCA, M1 segment), basilar artery (BA, pontine level), and bilateral vertebral arteries (VA, V4 segment). When stenosis was present, measurements were obtained from unaffected or minimally affected segments. Diameters were adjusted for intracranial volume (ICV) using residual correction. Bilateral ICA, MCA, and VA diameters were averaged to derive representative diameters for each vessel type. The mean composite intracranial artery diameter was calculated as the average of all seven arteries and standardized to a z score. BADE was further defined according to Smoker’s criteria, based on BA diameter, bifurcation height, and lateral displacement (Supplementary Methods and Figure S2).^20^

#### Whole and subregional brain segmentation

We registered all structural images to the T2W image space using FSL FLIRT (FMRIB Software Library, Oxford, United Kingdom). ICV was derived from brain extraction in the proton density image. Acute stroke lesions were manually delineated and excluded on the FLAIR image. White matter hyperintensities (WMH) and perivascular spaces (PVS) were segmented on FLAIR and T2W images, respectively, using a validated computational methods.^21, 22^ Co-registered FLAIR, T1W, and T2W images were used to generate the brain tissue mask using Gaussian clustering.

For vascular function, regions of interest (ROIs) were defined in normal-appearing white matter (NAWM), subcortical gray matter (SGM), and WMH. NAWM masks were generated using an in-house pipeline combining FreeSurfer (https://surfer.nmr.mgh.harvard.edu/) and FSL FAST outputs. The SGM mask was generated using FreeSurfer and included the caudate, putamen, pallidum, and thalamus.^21^ WMH, SGM, brainstem, and stroke lesions were excluded from the NAWM mask, and WMH and stroke lesions were excluded from the SGM mask. SGM and NAWM masks were eroded in T2W space by 1mm in all directions to reduce partial volume artifact. To minimize contamination from large blood vessels running along the inner ventricle surface, tissue adjacent to the ventricles was excluded using a ventricle mask dilated by 5mm laterally and by 4mm in the anterior, posterior, superior, and inferior directions. We then subtracted the dilated mask from the NAWM and WMH masks. Additional manual correction was applied to remove overlap with remaining large veins identified on SWI. All masks were visually inspected and manually edited when necessary to ensure accuracy.

#### Quantitative vascular function metrics

CVR, DCE-MRI, and PC-MRI data were processed for each ROI using validated techniques, as detailed in the Supplementary Methods.

#### cSVD markers

Baseline and 1-year cSVD markers were visually assessed according to STRIVE-2 criteria.^23^ A summary cSVD score was constructed as previously described.^24^ WMH volume was normalized to ICV and reported as %ICV, while PVS volume was normalized to the corresponding ROI (basal ganglia and centrum semiovale) and expressed as %ROIV. WMH progression was defined as the volume of new WMH developing from baseline NAWM during the 1-year follow-up. PVS growth was defined as the increase in PVS volume between baseline and 1 year. Incident infarcts were defined as new infarcts detected between index stroke and 1-year follow-up.^25^

### Statistical analysis

Descriptive data are presented as mean (standard deviation), median (interquartile range, IQR), or count (percentage). Between-group comparisons used *t* test or Mann-Whitney *U*test for continuous variables, and χ² test for categorical variables, as appropriate. PS and *v_p_* were rescaled (×10,000 and ×100, respectively) for comparability across variables. WMH (% ICV) and PVS (% ROIV) volumes were log_10_ transformed.

Primary outcomes were vascular function measures, including CVR, PS, *v_p_* in NAWM, WMH, and SGM, as well as ICA pulsatility index, superior sagittal sinus pulsatility index, and CSF stroke volume at the foramen magnum. Large artery characteristics (LAS, BADE, and arterial diameters) were treated as exposures. Multivariable linear regression was used to examine associations of BADE and LAS with vascular function, adjusting for age, sex, hypertension, diabetes mellitus, hyperlipidemia, smoking, and baseline mean arterial pressure, consistent with previous work.^8^ For arterial diameters, both mean composite diameter (z score derived from BA, bilateral ICAs, MCAs, and VAs) and vessel-specific diameters (mm) were analyzed. The optimal functional form was evaluated by comparing linear and restricted cubic spline (RCS) models using Akaike and Bayesian information criteria. To test whether associations varied across brain regions (NAWM, WMH, and SGM), we fitted mixed-effects models with a random intercept for participant ID and included region×diameter interaction terms. Post-hoc pairwise comparisons of slopes were conducted with Tukey adjustment.

Exploratory mediation analyses were performed for vascular function measures significantly associated with BADE, testing whether they mediated the relationship between dolichoectasia and cSVD markers. Outcomes included baseline summary cSVD score, number of lacunes and microbleeds, WMH and PVS volumes, as well as WMH progression, PVS growth, and incident infarcts over 1 year. Models were adjusted for age, sex, and vascular risk factors. Indirect effects and 95% confidence intervals (CI) were estimated using bootstrapping with 5,000 resamples, and the proportion mediated was calculated as the ratio of the indirect effect to the total effect. Sensitivity analyses tested the robustness of the associations between arterial diameters and vascular function by (i) substituting maximum instead of mean diameters for bilateral ICAs, MCAs, and VAs, and (ii) further adjusting for LAS.

All analyses were performed in R version 4.4.2 (R Foundation for Statistical Computing, Vienna, Austria). Given the exploratory and hypothesis-generating nature of the study, *P*-values were not adjusted for multiple comparisons, and findings should be interpreted as preliminary.

## Results

### Patient characteristics

A total of 224 participants were included (mean age 66.0 ± 11.2 years; 66.5% men). All had complete, analyzable structural MRI, whereas 42 CVR, 34 DCE-MRI, and 19 PC-MRI scans were not analyzable (Figure 1). There were no significant differences in age, sex, or vascular risk factors between participants with and without CVR or PC-MRI measurements. Participants with DCE-MRI were older (70.5 ± 9.3 vs 65.2 ± 11.3 years; *P*=0.004) and more likely to have a history of diabetes (41.2% vs 17.4%; *P*=0.004) compared with those without DCE-MRI (Supplementary Table S1).

At baseline, the median summary cSVD score was 2 (IQR 1-3), with a mean WMH volume of 0.94 ± 1.13 %ICV and mean total PVS volume of 4.19 ± 2.46 %ROIV (Table 1). Over 1 year, the volume of NAWM converting to WMH was 0.19 ± 0.23 %ICV, and there was an increase in PVS volume of 0.45 ± 1.66 %ROIV. BADE was identified in 36 participants (16.1%) and LAS in 46 (20.5%). Compared with those without BADE, participants with BADE were older, had larger intracranial arterial diameters (Supplementary Table S2), were more likely to present with an index lacunar stroke, and exhibited greater baseline cSVD burden. They also had more NAWM that progressed to WMH over 1 year. In unadjusted comparisons, BADE was associated with lower CVR in both NAWM (0.034 vs. 0.045 %/mmHg, *P*=0.003) and WMH (0.03 vs. 0.043 %/mmHg, *P*=0.033), while other vascular function measures did not differ. In contrast, participants with LAS more often presented with non-lacunar index strokes but did not differ from those without LAS in cSVD markers or any vascular functions (Table 1).

**Table 1.** Characteristics of the study population.

|  | Overall<br>(N=224) | BADE<br>(N=36) | Non-BADE<br>(N=188) | *P* value | LAS (N=46) | Non-LAS<br>(N=178) | *P* value |
| --- | --- | --- | --- | --- | --- | --- | --- |
| Age, y, mean ± SD | 66.0±11.2 | 69.7±10.9 | 65.2±11.1 | 0.021 | 69.8±9.74 | 65.0±11.3 | 0.011 |
| Sex, male, n (%) | 149 (66.5) | 28 (77.8) | 121 (64.4) | 0.171 | 28 (60.9) | 121 (68) | 0.462 |
| Index lacunar stroke, n (%) | 127 (56.7) | 29 (80.6) | 98 (52.1) | 0.003 | 19 (41.3) | 108 (60.7) | 0.028 |
| Baseline summary cSVD score, median (IQR) | 2 (1, 3) | 3 (1, 3.25) | 2 (0, 2) | <0.001 | 2 (1, 3) | 2 (1, 3) | 0.492 |
| Baseline WMH volume, %ICV, mean ± SD | 0.94±1.13 | 1.48±1.28 | 0.84±1.07 | <0.001 | 0.90±1.01 | 0.95±1.16 | 0.541 |
| Baseline PVS volume, %ROIV, mean ± SD | 4.19±2.46 | 5.55±2.71 | 3.93±2.32 | <0.001 | 4.15±2.01 | 4.21±2.57 | 0.684 |
| WMH progression at 1 year, %ICV, mean ± SD | 0.19±0.23 | 0.31±0.27 | 0.17±0.22 | <0.001 | 0.19±0.18 | 0.20±0.24 | 0.49 |
| Increase in PVS volume at 1 year, %ROIV, mean ± SD | 0.45±1.66 | 1.13±2.22 | 0.32±1.50 | 0.017 | 0.36±1.13 | 0.48±1.77 | 0.504 |
| CVR in NAWM, %/mm Hg, mean ± SD | 0.043±0.016 | 0.034±0.016 | 0.045±0.016 | 0.003 | 0.043±0.019 | 0.043±0.016 | 0.778 |
| CVR in WMH, %/mm Hg, mean ± SD | 0.041±0.043 | 0.03±0.037 | 0.043±0.044 | 0.033 | 0.034±0.044 | 0.042±0.043 | 0.293 |
| CVR in SGM, %/mm Hg, mean ± SD | 0.171±0.053 | 0.157±0.059 | 0.174±0.051 | 0.119 | 0.166±0.05 | 0.172±0.053 | 0.552 |
| PS in NAWM, 10<sup>-4</sup>min<sup>-1</sup>, mean ± SD | 0.17±1 | -0.03±1.18 | 0.21±0.96 | 0.277 | 0.19±0.82 | 0.17±1.05 | 0.852 |
| PS in WMH, 10<sup>-4</sup>min<sup>-1</sup>, mean ± SD | 0.87±1.62 | 0.85±1.56 | 0.87±1.63 | 0.805 | 0.93±1.23 | 0.85±1.72 | 0.429 |
| PS in SGM, 10<sup>-4</sup>min<sup>-1</sup>, mean ± SD | 0.77±1.27 | 0.70±1.60 | 0.78±1.20 | 0.911 | 0.69±0.97 | 0.79±1.35 | 0.371 |
| $v_p$ in NAWM, 10<sup>-2</sup>, mean ± SD | 0.64±0.19 | 0.63±0.21 | 0.64±0.18 | 0.697 | 0.67±0.21 | 0.63±0.18 | 0.176 |
| $v_p$ in WMH, 10<sup>-2</sup>, mean ± SD | 0.86±0.38 | 0.80±0.37 | 0.87±0.38 | 0.279 | 0.92±0.40 | 0.84±0.37 | 0.31 |
| $v_p$ in SGM, 10<sup>-2</sup>, mean ± SD | 1.49±0.36 | 1.44±0.35 | 1.50±0.36 | 0.423 | 1.54±0.37 | 1.48±0.35 | 0.325 |
| Internal carotid artery pulsatility index, mean ± SD | 1.16±0.36 | 1.23±0.33 | 1.15±0.36 | 0.075 | 1.15±0.40 | 1.16±0.35 | 0.804 |
| Superior sagittal sinus pulsatility index, mean ± SD | 0.53±0.22 | 0.56±0.22 | 0.52±0.22 | 0.416 | 0.56±0.25 | 0.52±0.21 | 0.726 |
| CSF stroke volume at the foramen magnum, ml, mean ± SD | 0.55±0.30 | 0.57±0.29 | 0.55±0.30 | 0.733 | 0.49±0.28 | 0.57±0.30 | 0.101 |

Abbreviations: BADE, basilar artery dolichoectasia; LAS, large artery stenosis; SD, standard deviation; IQR, interquartile range; cSVD, cerebral small vessel disease; WMH, white matter hyperintensity; ICV, intracranial volume; PVS, perivascular space; ROIV, volume of region of interest; CVR, cerebrovascular reactivity; NAWM, normal appearing white matter; SGM, subcortical gray matter; PS, permeability surface area product; $v_p$ , plasma volume; CSF, cerebrospinal fluid.

### BADE, LAS and cerebrovascular function

In multivariable regressions adjusting for age, sex, and vascular risk factors, BADE was independently associated with lower CVR in NAWM (β -0.010, 95% CI -0.016 to -0.004; *P*=0.003) (Table 2). No significant associations were observed between BADE and CVR in WMH or SGM, nor with PS, *v_p_*, or pulsatility. LAS was not associated with CVR, PS, or pulsatility, but was related to higher *v_p_*in WMH (β 0.138, 95% CI 0.007-0.269; *P*=0.04). When individual arterial stenosis was examined (Supplementary Table S3), ICA stenosis (*n*=17) was associated with lower ICA pulsatility index (β -0.174, 95% CI -0.341 to -0.007; *P*=0.041), while VA stenosis (*n*=11) was associated with higher superior sagittal sinus pulsatility index (β 0.165, 95% CI 0.050-0.281; *P*=0.005).

**Table 2.** Associations of BADE and LAS with cerebrovascular function.

| Exposure Variable | Basilar artery dolichoectasia<sup>a</sup> |  | Large artery stenosis<sup>a</sup> |  |
| --- | --- | --- | --- | --- |
| | $\beta$ (95% CI) | *P* value | $\beta$ (95% CI) | *P* value |
| **Cerebrovascular reactivity, %/mm Hg** |  |  |  |  |
| CVR in normal appearing white matter | -0.01 (-0.016, -0.004) | 0.003 | 0.001 (-0.005, 0.007) | 0.824 |
| CVR in white matter hyperintensity | -0.011 (-0.028, 0.007) | 0.215 | -0.008 (-0.024, 0.009) | 0.359 |
| CVR in subcortical gray matter | -0.012 (-0.033, 0.009) | 0.247 | -0.004 (-0.023, 0.016) | 0.711 |
| **Permeability surface area product, 10<sup>-4</sup>min<sup>-1</sup>** |  |  |  |  |
| PS in normal appearing white matter | -0.181 (-0.580, 0.219) | 0.373 | -0.003 (-0.359, 0.353) | 0.987 |
| PS in white matter hyperintensity | -0.078 (-0.728, 0.572) | 0.813 | 0.004 (-0.574, 0.582) | 0.99 |
| PS in subcortical gray matter | -0.241 (-0.746, 0.265) | 0.349 | -0.056 (-0.507, 0.395) | 0.806 |
| **Plasma volume, 10<sup>-2</sup>** |  |  |  |  |
| $v_p$ in normal appearing white matter | 0.003 (-0.070, 0.077) | 0.935 | 0.034 (-0.032, 0.099) | 0.31 |
| $v_p$ in white matter hyperintensity | -0.030 (-0.179, 0.119) | 0.694 | 0.138 (0.007, 0.269) | 0.04 |
| $v_p$ in subcortical gray matter | -0.021 (-0.161, 0.118) | 0.764 | 0.037 (-0.087, 0.161) | 0.556 |
| **Phase contrast MRI** |  |  |  |  |
| Internal carotid artery pulsatility index | 0.001 (-0.127, 0.129) | 0.985 | -0.043 (-0.158, 0.072) | 0.465 |
| Superior sagittal sinus pulsatility index | -0.011 (-0.086, 0.063) | 0.764 | 0.001 (-0.065, 0.067) | 0.982 |
| CSF stroke volume at the foramen magnum, ml | 0.027 (-0.093, 0.147) | 0.66 | -0.064 (-0.17, 0.043) | 0.239 |

<sup>a</sup>Models were adjusted for age, sex, hypertension, diabetes mellitus, hyperlipidemia, smoking, and mean arterial pressure. Abbreviations: CI, confidential interval; CVR, cerebrovascular reactivity; PS, permeability surface area product; $v_p$ , plasma volume; CSF, cerebrospinal fluid.

### Arterial diameters and cerebrovascular function

Model comparisons showed that linear models best captured associations of arterial diameters with CVR, *v_p_*, and pulsatility, while RCS models provided a better fit for PS (Supplementary Table S4).

Larger mean composite intracranial arterial diameter was associated with lower CVR in NAWM (β -0.0067, 95% CI -0.0115 to -0.0020; *P*=0.006) and WMH (β -0.0146, 95% CI -0.0277 to -0.0015; *P*=0.030) (Table 3, Figure 2). In mixed-effects models, the overall region×diameter interaction across NAWM, WMH, and SGM was not significant (χ²=2.73, df=2, *P*=0.26). Pairwise comparisons confirmed no slope differences among regions (Supplementary Table S5, Figure S3), indicating that the diameter-CVR relationship was consistent across tissue types. In vessel-specific analyses, larger MCA diameter was consistently related to lower CVR in NAWM (β -0.0087, 95% CI -0.0169 to -0.0004; *P*=0.04), WMH (β -0.0269, 95% CI -0.0491 to -0.0047; *P*=0.018), and SGM (β -0.0264, 95% CI -0.0525 to -0.0003; *P*=0.048). Other arterial diameters also tended to show similar negative associations with CVR, although these did not reach statistical significance.

**Figure 2.**
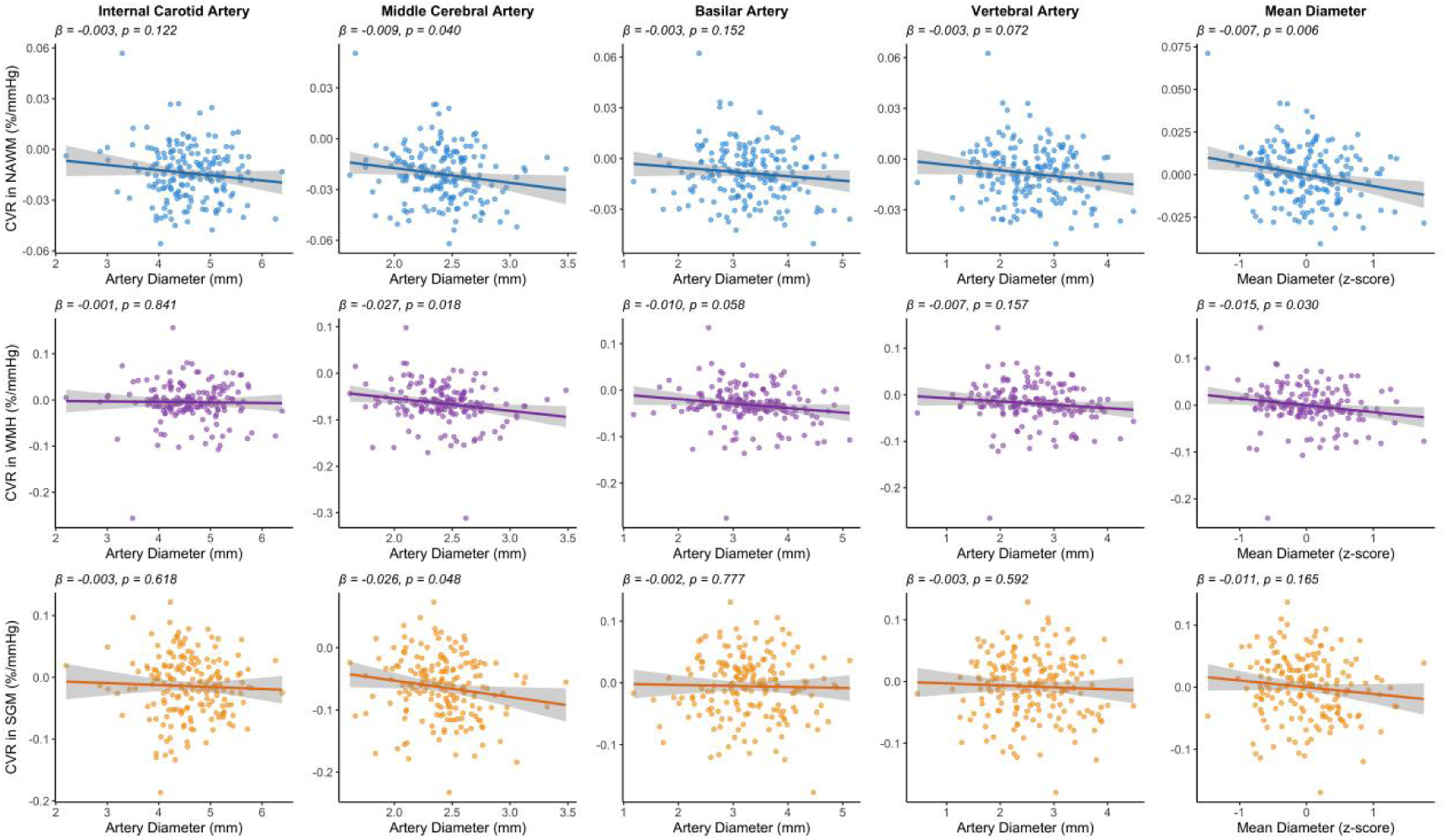
Scatter plots with fitted regression lines showing associations of intracranial arterial diameters with cerebrovascular reactivity in NAWM (blue), WMH (red), and SGM (green). Models were adjusted for age, sex, hypertension, diabetes mellitus, hyperlipidemia, smoking, and mean arterial pressure. Abbreviations: NAWM, normal-appearing white matter; WMH, white matter hyperintensity; SGM, subcortical gray matter; CVR, cerebrovascular reactivity.

**Table 3.** Linear regressions of intracranial arterial diameters with cerebrovascular reactivity.

|  | CVR in NAWM<sup>a</sup> |  | CVR in WMH<sup>a</sup> |  | CVR in SGM<sup>a</sup> |  |
| --- | --- | --- | --- | --- | --- | --- |
| | $\beta$ (95% CI) | *P* value | $\beta$ (95% CI) | *P* value | $\beta$ (95% CI) | *P* value |
| Mean diameter | -0.0067 (-0.0115, -0.0020) | 0.006 | -0.0146 (-0.0277, -0.0015) | 0.030 | -0.0109 (-0.0262, 0.0045) | 0.165 |
| ICA diameter | -0.0031 (-0.0070, 0.0008) | 0.122 | -0.0011 (-0.0117, 0.0095) | 0.841 | -0.0031 (-0.0156, 0.0093) | 0.618 |
| MCA diameter | -0.0087 (-0.0169, -0.0004) | 0.040 | -0.0269 (-0.0491, -0.0047) | 0.018 | -0.0264 (-0.0525, -0.0003) | 0.048 |
| BA diameter | -0.0026 (-0.0062, 0.0010) | 0.152 | -0.0096 (-0.0196, 0.0003) | 0.058 | -0.0016 (-0.0130, 0.0098) | 0.777 |
| VA diameter | -0.0033 (-0.0070, 0.0003) | 0.072 | -0.0072 (-0.0171, 0.0028) | 0.157 | -0.0032 (-0.0148, 0.0085) | 0.592 |

<sup>a</sup> Models were adjusted for age, sex, hypertension, diabetes mellitus, hyperlipidemia, smoking, and mean arterial pressure. Mean diameter was standardized, so $\beta$ values represent change in outcome per 1-SD larger mean diameter; vessel-specific diameters were analyzed in mm. Abbreviations: CI, confidential interval; NAWM, normal-appearing white matter; WMH, white matter hyperintensity; SGM, subcortical gray matter; ICA, internal carotid artery; MCA, middle cerebral artery; BA, basilar artery; VA, vertebral artery; CVR, cerebrovascular reactivity.

A U-shaped relationship was observed between arterial diameter and PS (Supplementary Table S6, Figure 3). Both smaller and larger mean diameters were associated with higher PS, with significant non-linear (U-shaped) associations in NAWM (*P*<0.001) and WMH (*P*=0.024). In spline mixed-effects models, the region×diameter interaction was not significant (χ²=1.12, df=4, *P*=0.89) (Supplementary Table S5, Figure S3), indicating that the U-shaped association was consistent across different regions. Vessel-specific analyses showed similar U-shaped relationships for MCA diameter in NAWM (*P*<0.001), WMH (*P*=0.049), and SGM (*P*=0.045), and for BA diameter in NAWM (P=0.032) and SGM (P=0.046). Neither mean or vessel-specific diameters were associated with *v_p_* in NAWM, WMH, or SGM (Supplementary Table S7).

**Figure 3.**
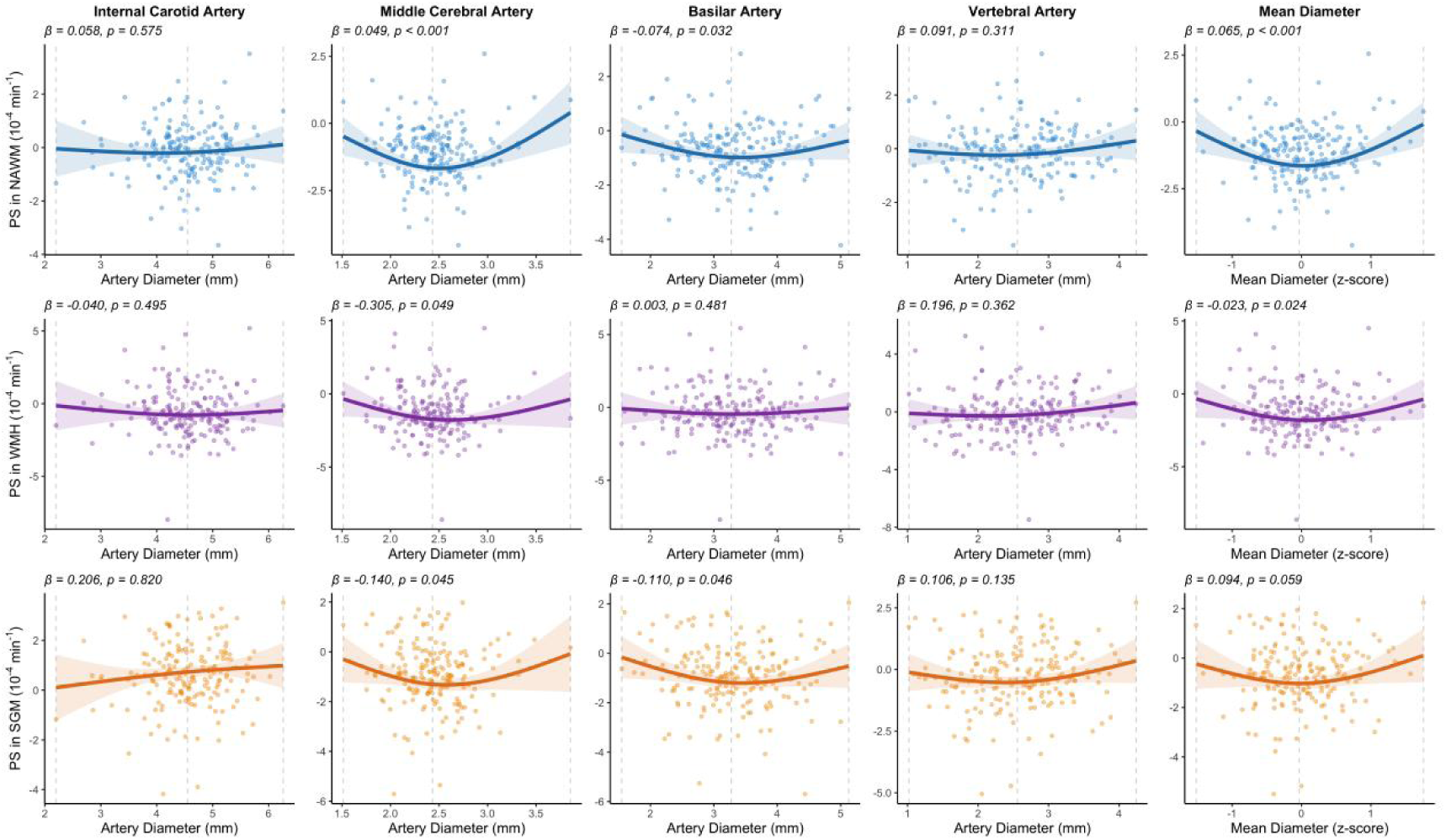
Scatter plots with restricted cubic spline fits showing U-shaped associations between intracranial arterial diameters and permeability surface area product in NAWM (blue), WMH (red), and SGM (green). Models were adjusted for age, sex, hypertension, diabetes mellitus, hyperlipidemia, smoking, and mean arterial pressure. Abbreviations: NAWM, normal-appearing white matter; WMH, white matter hyperintensity; SGM, subcortical gray matter; PS, permeability surface area product.

For pulsatility measures, no significant associations were found between arterial diameters and arterial or venous sinus pulsatility indices (Supplementary Table S8). Larger MCA diameter was associated with higher CSF stroke volume at the foramen magnum (β=0.153, 95% CI 0.016-0.29; *P*=0.029), with similar but weaker trends for ICA diameter and mean composite arterial diameter.

### Mediation analysis

Given the observed associations between BADE and cSVD markers, mediation analyses were performed with BADE as the exposure, CVR in NAWM as the mediator, and cSVD markers at baseline and their 1-year progression as the outcomes. CVR significantly mediated the associations of BADE with summary cSVD score, number of lacunes and microbleeds at baseline, and WMH and PVS progression over 1 year (Figure 4A, Supplementary Table S9). Specifically, for NAWM that progressed to WMH at 1 year, CVR accounted for 19.9% (95% CI 3.9-55.9%; *P*=0.011) of the total effect, with an indirect effect of 0.053 (95% CI 0.009-0.116; *P*=0.007) (Figure 4B).

**Figure 4.**
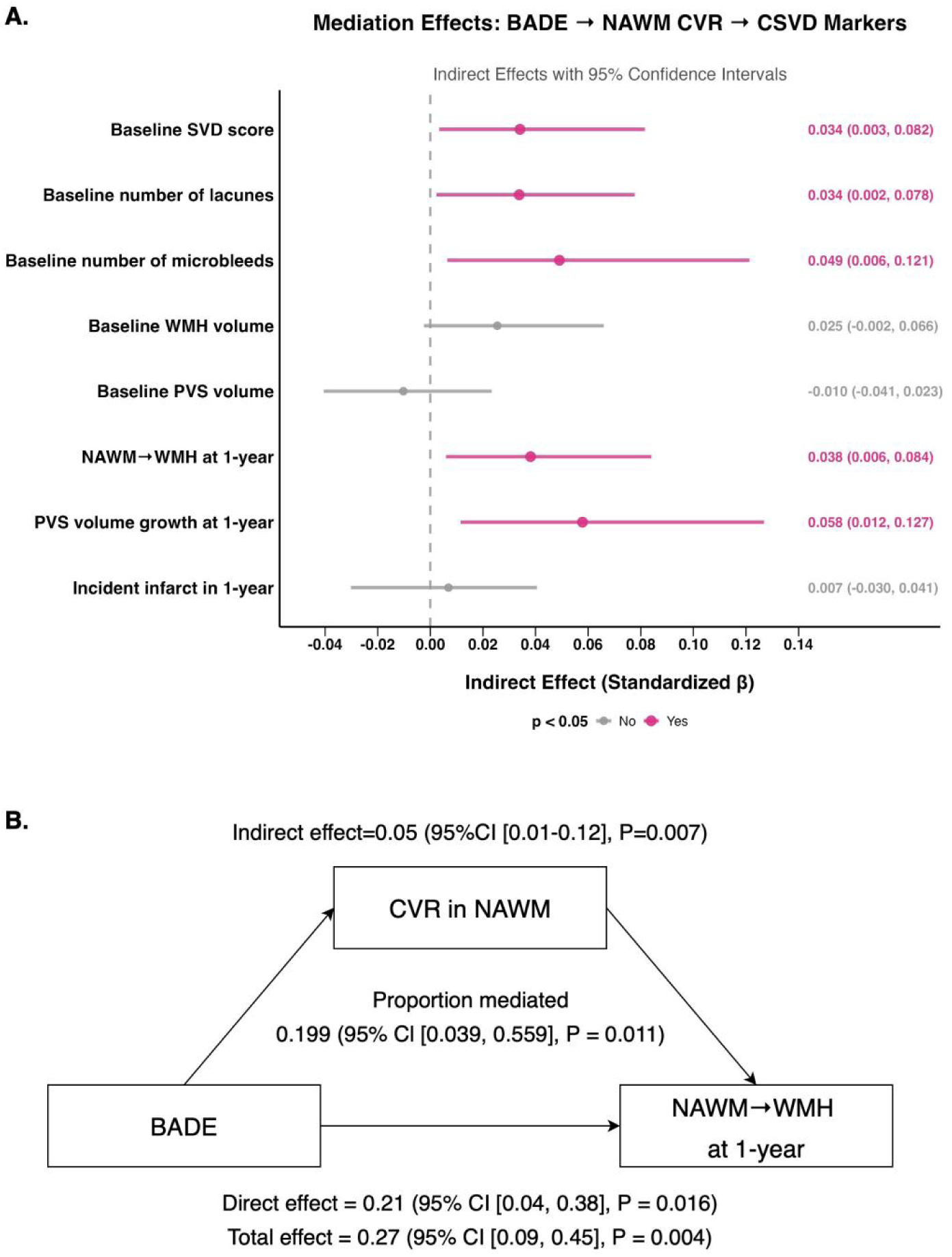
Mediation effects of CVR in NAWM on the associations between BADE and cSVD markers. (A) Forest plot of indirect effects. (B) Path diagram illustrating the mediation model for WMH progression at 1 year. All models were adjusted for age, sex, and vascular risk factors. Abbreviations: BADE, basilar artery dolichoectasia; CSVD, cerebral small vessel disease; CVR, cerebrovascular reactivity; NAWM, normal appearing white matter; WMH, white matter hyperintensity; PVS, perivascular space.

### Sensitivity analyses

Findings were robust across sensitivity analyses. Substituting maximum instead of mean diameters of bilateral MCA, ICA, and VA yielded similar associations with CVR and PS in NAWM, WMH, and SGM (Supplementary Table S10). Additional adjustment for LAS did not alter the results (Supplementary Table S11).

## Discussion

In this prospective cohort of lacunar stroke enriched for cSVD features, with a comparator group of similarly mild cortical stroke, we applied multimodal MRI to investigate the associations between large artery phenotypes and several measures of cerebrovascular function. Intracranial arterial dilatation, most notably MCA dilatation, was associated with lower CVR, higher CSF stroke volume, and a U-shaped relationship with BBB permeability. Lower NAWM CVR partially mediated the effect of BADE on 1-year progression of WMH and PVS volumes. In contrast, LAS was unrelated to CVR or BBB permeability but linked to higher plasma volume. Overall, these findings indicate that distinct large artery phenotypes coexist with small-vessel dysfunction and altered intracranial hemodynamics, which may help identify individuals at risk of accelerated cSVD progression.

Evidence on the relationship between intracranial dolichoectasia and CVR is limited. We found that arterial dilatation was associated with lower CVR, likely reflecting both structural and geometric changes that reduce compliance. Structurally, dolichoectatic arteries exhibit medial degeneration with elastin loss, collagen accumulation, and smooth muscle disruption, compromising wall elasticity.^1, 26^ Geometrically, larger diameters reduce the relative capacity of the wall to expand with each pulse. These alterations weaken the buffering function of large arteries, may transmit abnormal hemodynamic loads to penetrating arterioles, and ultimately impair autoregulation.^27^ Binary BADE classification was associated only with NAWM CVR, suggesting stage-dependent vulnerability. Compared with WMH, NAWM retains more functional responsiveness and is more sensitive to upstream hemodynamic stress, whereas WMH represents chronically more injured tissue where a floor effect may obscure further associations, as suggested by visibly dilated small vessels in severe WMH, where CVR is particularly poor and tissue is cavitating.^28^ Notably, our longitudinal analyses demonstrated that reduced NAWM CVR partially mediated the association between BADE and progression of WMH and PVS. Consistent with Fisher’s^29^ pathological descriptions of intrinsic microarteriolar dilatation and medial hyalinization in cSVD, widening at the large-artery level may therefore represent a macroscopic ‘signal’ of underlying microvascular pathology and exhausting or exhausted vasodilatory capacity.^3^ Collectively, these findings reinforce dolichoectasia as a marker of decompensated vascular dysfunction and highlight NAWM CVR as a potentially modifiable target to slow disease progression.

Emerging genetic evidence further supports this interpretation. Genetic studies of WMH have implicated variants in genes encoding vascular structural proteins, such as collagen type IV (*COL4A1/COL4A2*), and in regulators of extracellular matrix homeostasis such as *HTRA1*.^30–34^ In addition, investigations of vascular traits have demonstrated shared genetic determinants between reduced aortic compliance and WMH, supporting a common vulnerability of both large and small artery walls.^35^ Many of these loci converge on pathways involving extracellular matrix organization, vascular smooth muscle contractility, and vessel wall remodeling, indicating that impaired integrity and compliance are genetically influenced traits across the vascular tree that predispose to cSVD.

Arterial diameter showed a U-shaped association with BBB permeability, with both small and large calibers linked to greater leakage. At the lower end, smaller arteries may reduce downstream perfusion and shear stress, predisposing to endothelial injury and impaired energy-dependent transport, thereby compromising barrier integrity. Consistent with this, studies in cSVD have reported that regions of lower cerebral blood flow tend to show greater BBB leakage.^36^ At the upper end, arterial dilatation occurs in parallel with microvessel widening, which has been associated with neurovascular unit dysfunction and BBB disruption in cSVD. Thus, rather than reflecting a single pathway, BBB integrity may be compromised at both extremes of vascular caliber, underscoring its vulnerability to hemodynamic stress, although the mechanisms require further clarification.

We found no associations between arterial or venous pulsatility and arterial widening. Although still controversial, longitudinal data indicate that baseline pulsatility does not predict cSVD progression, whereas baseline WMH and PVS volumes predict subsequent increases in pulsatility.^12, 37^ Additional evidence suggests that the relation between arterial pulsatility and WMH may reflect shared upstream processes such as arterial stiffening and falling diastolic pressure.^38^ Taken together, these findings indicate that large artery dilatation and cSVD represent related structural changes, whereas increased pulsatility is more likely a consequence or coexisting phenomenon of microvascular damage rather than a causal driver. Prior studies examining CSF pulsatility in cSVD have yielded mixed results.^12, 14, 39^ In our study, however, MCA widening was associated with greater CSF stroke volume, suggesting altered coupling between large arteries, intracranial fluid dynamics and interstitial homeostasis, although mechanisms and causal direction remain uncertain.

The absence of any relationship between LAS and CVR, PS, or pulsatility supports the view that the cSVD is not atheromatous or simply due to ‘hypoperfusion’ resulting from the stenosis. Instead, LAS was associated with higher plasma volume fraction in WMH, which may reflect a compensatory increase in cerebral blood volume to preserve flow within autoregulatory limits distal to stenosis.

This study has several important strengths. To our knowledge, it is the first to systematically investigate the vascular functional consequences of large-artery phenotypes and cerebral microvasculature in vivo. By combining binary classifications with continuous diameter measures, we captured both overt and subclinical vascular remodeling. The use of a prospective, well-characterized cohort with controlled stroke subtype, standardized stroke prevention medications, multimodal MRI, and blinded image analysis provided a robust platform for these assessments. Several limitations should also be acknowledged. Vascular function was measured at baseline, so the cross-sectional design precludes definitive causal inference, and mediation analyses should be interpreted as exploratory. Longitudinal studies are needed to determine the temporal associations between large artery pathology and vascular function. PC-MRI pulsatility measures have limited spatial and temporal resolution, which may underestimate subtle effects. Finally, missing, or unanalyzable vascular functional MRI data reduced the effective sample size, potentially limiting statistical power.

## Conclusion

Large artery dilatation may be regarded as a macroscopic signal of small-vessel dysfunction. It was associated with lower CVR, a U-shaped relationship with BBB permeability, and greater CSF pulsatility. Reduced CVR in NAWM partially mediated the impact of dolichoectasia on cSVD progression. By contrast, LAS was linked to higher plasma volume in WMH. These findings demonstrate that large-artery phenotypes are integrally related to cSVD-related brain injury perhaps through different hemodynamic pathways and identify NAWM CVR as a potential therapeutic target to modify vascular function and slow structural progression.

## Data Availability

All data produced in the present study are available upon reasonable request to the authors

