## Supplemental Material for "Large artery phenotypes, cerebrovascular function, and progression of cerebral small vessel disease"

### Supplementary Materials


1. **Supplementary Methods**

1.1 Image acquisition

1.2 Image analysis

1.2.1 Large artery stenosis assessment

1.2.2 Supplementary Figure 1. Illustration of severe stenosis in the left middle cerebral artery

1.2.3 Intracranial arterial dolichoectasia assessment

1.2.4 Supplementary Figure 2. Assessment of intracranial artery dolichoectasia

1.2.5 CVR

1.2.6 DCE-MRI

1.2.7 Phase contrast-MRI

2. **Supplementary Results**

2.1 Supplementary Table 1. Comparison of baseline characteristics between participants with and without CVR, DCE-MRI, and PC-MRI measurements

2.2 Supplementary Table 2. Distribution of intracranial artery diameters

2.3 Supplementary Table 3. Associations of ICA, MCA, VA, and BA stenosis with cerebrovascular function

2.4 Supplementary Table 4. Model comparison of linear vs. restricted cubic spline regressions across exposure–outcome pairs

2.5 Supplementary Table 5. Model fit statistics comparing models with and without the region×diameter interaction

2.6 Supplementary Figure 3. Associations between mean arterial diameter and cerebrovascular function across regions.

2.7 Supplementary Table 6. Linear regressions of intracranial arterial diameters with plasma volume

2.8 Supplementary Table 7. Linear regressions of intracranial arterial diameters with pulsatility

2.9 Supplementary Table 8. Mediation analyses of the associations between basilar artery dolichoectasia (exposure) and cerebral small vessel disease markers (outcome), with cerebrovascular reactivity in normal-appearing white matter as the mediator

2.10 Supplementary Table 9. Sensitivity analysis using maximum instead of mean diameters in relation to cerebrovascular function

2.11 Supplementary Table 10. Sensitivity analysis additionally adjusting for large artery stenosis in the associations of intracranial arterial diameters with cerebrovascular function

#### 1. Supplementary Methods

##### 1.1 Image acquisition

We performed a 2D gradient-echo echo-planar imaging scan to measure CVR (TR/TE=1550/30ms; flip angle, 67°; isotropic resolution, 2.5mm$^3$). During the 12-minute CVR scan we administered medical air and 6% $CO_2$ enriched air ($CO_2$: O2:N2, 6%:21%:73%) alternately for 2 and 3 minutes, respectively. Patients wore a carefully fitted anaesthetic face mask attached to a unidirectional open breathing circuit (Intersurgical, Wokingham, United Kingdom). We monitored other physiological parameters: end-tidal $CO_2$, end-tidal $O_2$, oxygen saturation level, and heart and respiration rates.

We acquired DCE-MRI using a 3D sagittal T1W sGRE with 32 volumes acquired at a temporal resolution of 39.6s. After acquiring three pre-contrast volumes, we intravenously injected a dose of 0.1mmol/kg body weight gadobutrol (1M Gadovist, Bayer AG, Leverkusen, Germany) delivered over a period of 110-130s using a power injector followed by a 20ml saline flush.

We performed 3 separate 2D PC-MRI acquisitions to assess carotid arteries, venous sinuses, and CSF spaces: one axial slice perpendicular to the external carotid arteries and ICAs at the spine C2-3 level (TR/TE=19.6/5.8ms; flip angle, 12°; spatial resolution, 1.0×1.0 mm$^2$; temporal resolution, 39.2ms; and venc=70cm/s), one coronal slice bisecting the superior sagittal sinus, straight sinus, and transverse sinuses (TR/TE=21.7/6.6ms; flip angle, 12°; spatial resolution, 0.71×0.71mm$^2$; temporal resolution, 43.4ms; and $v_{enc}$=50cm/s), and one axial slice perpendicular to the C2-3 level spinal cord (TR/TE=25.2/8.5ms; flip angle, 12°; spatial resolution, 0.83×0.83 mm$^2$; temporal resolution, 50.4ms; and $v_{enc}$=6cm/s). We interpolated the phase images across 32 timeframes, covering the cardiac cycle, using retrospective cardiac gating, a well-established multicenter study and trial method.

##### 1.2 Image analysis

###### 1.2.1 Large artery stenosis assessment

As MR angiography was not routinely performed in MSS3, Intracranial artery stenosis was assessed using DCE-MRI. Images were reviewed in sagittal, axial, and coronal planes, selecting the time point with optimal arterial enhancement for lumen visualization. Stenosis was graded at the point of maximal narrowing of the contrast-filled lumen using established criteria. Intracranial arteries assessed included internal carotid arteries (ICA), middle cerebral arteries (MCA), anterior cerebral arteries, posterior cerebral arteries, vertebral arteries (VA), and the basilar artery (BA). When stenosis was suspected, conventional structural sequences were reviewed for confirmation (Supplementary Fig. 1). The cervical ICA was assessed at presentation of the index stroke using carotid ultrasound, and, where available, CT angiography or MR angiography. Stenosis was classified according to the North American Symptomatic Carotid Endarterectomy Trial (NASCET) criteria. Large artery stenosis (LAS) was defined as the presence of ≥50% stenosis of any intracranial artery and cervical ICA stenosis.

**1.2.2 Supplementary Figure 1. Illustration of severe stenosis in the left middle cerebral artery.** Panels A-D show consecutive sagittal slices from a dynamic contrast-enhanced MRI scan, progressing from the central (A) to peripheral slices (D), with red arrows indicating the site of stenosis. Panels E and F display axial T2-weighted images of the same artery.

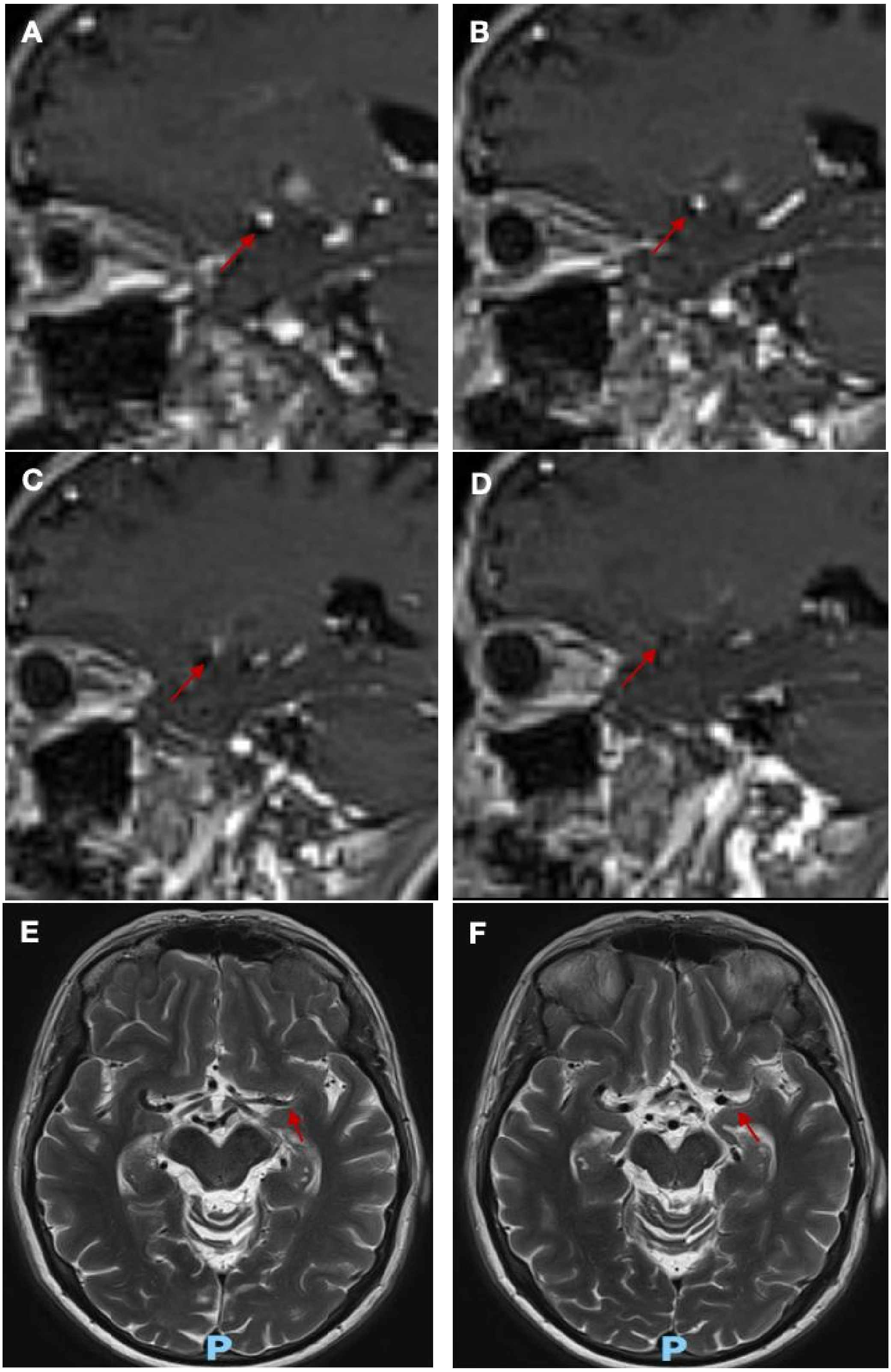

##### 1.2.3 Intracranial arterial dolichoectasia assessment

Arterial diameters were measured on axial T2W images (Supplementary Fig. 2). We assessed diameters of seven intracranial arteries: the BA at the midpontine level, bilateral VAs at the V4 segment, bilateral ICAs at the cavernous segment, and bilateral MCAs at the M1 segment. When stenosis was present, measurements were obtained from unaffected or minimally affected segments. Given ICV is a determinant of intracranial artery caliber, arterial diameters were adjusted for ICV using residual correction methods. Bilateral ICA, MCA, and VA values were averaged to derive representative diameters for each vessel type. The mean intracranial artery diameter was calculated as the average of these seven arteries and standardized to a z score. For basilar artery evaluation, we additionally assessed bifurcation height and lateral displacement using a validated four-point scoring system (Figure S2). Bifurcation height categories included: 0=at/below dorsum sellae, 1=suprasellar cistern, 2=third ventricle floor level, 3=elevating third ventricle floor. Lateral displacement was graded as: 0=midline, 1=medial to clivus/dorsum sellae margin, 2=lateral to this margin, 3=cerebellopontine angle cistern. BADE was defined according to Smoker criteria by meeting any criterion: BA diameter >4.5 mm, bifurcation height score ≥2, or lateral displacement score ≥2.

##### 1.2.4 Supplementary Figure 2. Assessment of intracranial artery dolichoectasia.

(A) Measurement of the bilateral internal carotid artery diameters at the vertical cavernous segment, and the maximum diameter of the basilar artery on axial T2-weighted imaging. (B) Measurement of the bilateral middle cerebral artery diameters at the M1 segment on axial T2-weighted imaging. (C) Measurement of the bilateral vertebral artery diameters at the V4 segment on axial T2-weighted imaging. (D) Grading of basilar artery of lateral displacement using a validated 4-point grading scale (0–3). (E) Grading of basilar artery bifurcation height in the sagittal plane on T1-weighted imaging (scale 0-3).

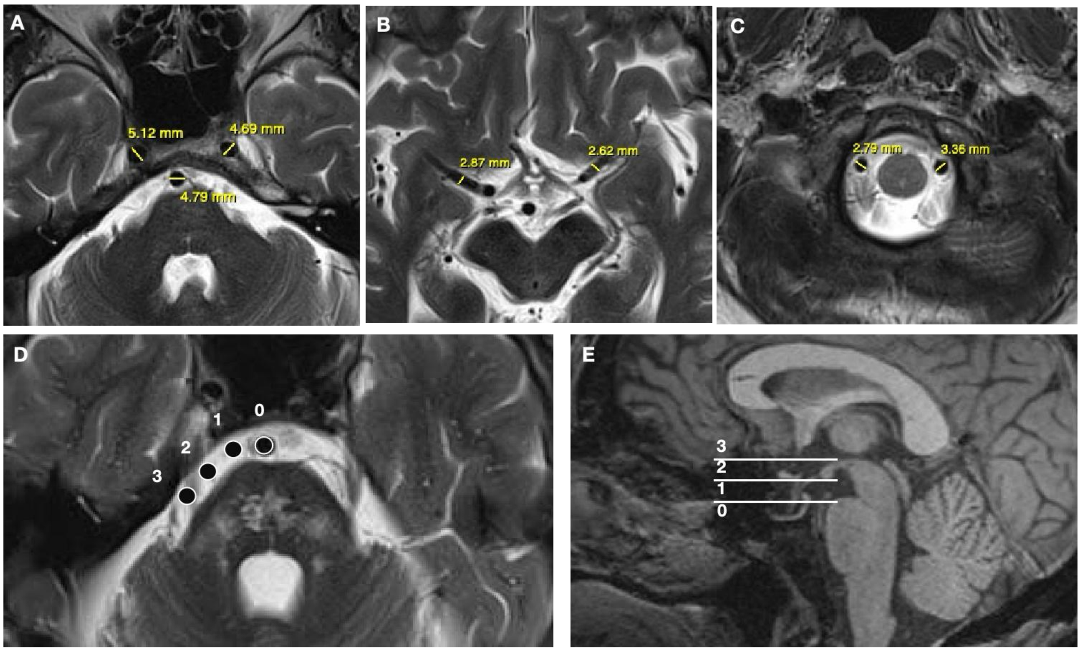

##### 1.2.5 CVR

CVR data analysis was performed by one investigator and independently verified by a second researcher. We extracted CVR measurements from three distinct tissue compartments: NAWM, WMH, and SGM. Following temporal realignment of BOLD sequences, tissue masks were transformed to the mean BOLD space using FSL FLIRT registration. Linear regression modeling was applied to extract mean BOLD signal from each region of interest, incorporating time-shifted end-tidal $CO_2$ profiles and linear drift correction (volume number) as regressors. The optimal temporal delay was determined by minimizing sum of squared residuals. CVR was calculated as the regression coefficient for end-tidal $CO_2$ divided by baseline BOLD signal (mean intensity during initial 30 volumes of room air) and multiplied by 100, yielding relative signal change per unit $CO_2$ change expressed as %/mmHg.

##### 1.2.6 DCE-MRI

We spatially realigned the brain extracted DCE-MRI images using SPM12. Per consensus guidelines we estimated a vascular input function by manually selecting five voxels in the SSS and taking the mean value at each time point using an established method. We registered the T2W scan to the mean pre-contrast image using FSL FLIRT and transformed the tissue masks into the DCE space. We calculated the signal enhancement for each volume relative to the mean pre-contrast signal before estimating the Patlak tracer kinetic parameters, permeability surface area (PS) and blood plasma volume ($v_p$), assuming the no-exchange limit for water transport across the BBB and corrected for $B_1^+$ error.

##### 1.2.7 Phase contrast-MRI

We calculated the maximum PC-MRI signal magnitude across the cardiac cycle per pixel to help with vessel identification and drew ROIs for the ICAs, vertebral arteries, internal jugular veins, superior sagittal sinus, straight sinus, and transverse sinuses lumens and subarachnoid CSF space at the foramen magnum using established methods. Using carefully positioned background ROIs placed near to the vessels in regions with no visible flow we corrected for background phase error. For each vessel we summed the total flow across the cardiac cycle and calculated the pulsatility index based on an adapted version of Gosling's equation. We calculated subarachnoid CSF stroke volume by averaging the absolute flow volume in caudal and cranial directions.

#### 2. Supplementary Results

##### 2.1 Supplementary Table 1. Comparison of baseline characteristics between participants with and without CVR, DCE-MRI, and PC-MRI measurements

| Variable | Without CVR (n=42) | With CVR (n=182) | *P* value | Without DCE-MRI (n=34) | With DCE-MRI (n=190) | *P* value | Without PC-MRI (n=19) | With PC-MRI (n=205) | *P* value |
|---|---|---|---|---|---|---|---|---|---|
| Age, y, mean ± SD | 64.5 ± 11.1 | 66.3 ± 11.2 | 0.351 | 70.5 ± 9.3 | 65.2 ± 11.3 | 0.004 | 61.6 ± 10.8 | 66.4 ± 11.2 | 0.074 |
| Sex, male, n (%) | 26 (61.9%) | 123 (67.6%) | 0.602 | 22 (64.7%) | 127 (66.8%) | 0.963 | 13 (65.0%) | 136 (66.7%) | 1 |
| Smoker: current or ex<1y, n (%) | 10 (23.8%) | 30 (16.5%) | 0.371 | 6 (17.6%) | 34 (17.9%) | 1 | 5 (25.0%) | 35 (17.2%) | 0.57 |
| Hypertension, n (%) | 26 (61.9%) | 128 (70.3%) | 0.380 | 26 (76.5%) | 128 (67.4%) | 0.393 | 13 (65.0%) | 141 (69.1%) | 0.899 |
| Diabetes mellitus, n (%) | 11 (26.2%) | 36 (19.8%) | 0.478 | 14 (41.2%) | 33 (17.4%) | 0.004 | 4 (20.0%) | 43 (21.1%) | 1 |
| Hyperlipidemia, n (%) | 32 (76.2%) | 134 (73.6%) | 0.883 | 26 (76.5%) | 140 (73.7%) | 0.897 | 17 (85.0%) | 149 (73.0%) | 0.296 |
| Prior history of stroke or TIA, n (%) | 9 (21.4%) | 27 (14.8%) | 0.415 | 8 (23.5%) | 28 (14.7%) | 0.302 | 2 (10.0%) | 34 (16.7%) | 0.749 |

Abbreviations: SD, standard deviation; TIA, transient ischemic attack; CVR, cerebrovascular reactivity; DCE, dynamic contrast-enhanced; PC, phase contrast.

##### 2.2 Supplementary Table 2. Distribution of intracranial artery diameters

| | Overall (N=224) | BADE (N=36) | Non-BADE (N=188) | *P* value | LAS (N=46) | Non-LAS (N=178) | *P* value |
|---|---|---|---|---|---|---|---|
| Internal carotid artery, mm, mean ± SD | 4.57±0.64 | 4.81±0.76 | 4.53±0.61 | 0.052 | 4.49±0.73 | 4.60±0.62 | 0.23 |
| Middle cerebral artery, mm, mean ± SD | 2.44±0.33 | 2.64±0.42 | 2.40±0.3 | <0.001 | 2.45±0.35 | 2.43±0.33 | 0.798 |
| Basilar artery, mm, mean ± SD | 3.27±0.72 | 3.93±0.63 | 3.14±0.66 | <0.001 | 3.27±0.68 | 3.27±0.73 | 0.758 |
| Vertebral artery, mm, mean ± SD | 2.57±0.69 | 3.07±0.65 | 2.47±0.66 | <0.001 | 2.69±0.64 | 2.54±0.71 | 0.225 |
| Mean artery diameter (z-score), mean ± SD | 0±0.56 | 0.51±0.62 | -0.10±0.49 | <0.001 | 0.013±0.59 | -0.003±0.55 | 0.897 |

Abbreviations: BADE, basilar artery dolichoectasia; LAS, large artery stenosis; SD, standard deviation.

#### 2.3 Supplementary Table 3. Associations of ICA, MCA, VA, and BA stenosis with cerebrovascular function

| | ICA stenosis (N=17) | | MCA stenosis (N=14) | | VA stenosis (N=11) | | BA stenosis (N=5) | |
|---|---|---|---|---|---|---|---|---|
| Exposure Variable | β (95% CI) | P value | β (95% CI) | P value | β (95% CI) | P value | β (95% CI) | P value |
| **Cerebrovascular reactivity, %/mm Hg** | | | | | | | | |
| CVR in normal appearing white matter | 0.003 (-0.006, 0.013) | 0.515 | 0.004 (-0.006, 0.013) | 0.479 | 0.001 (-0.01, 0.012) | 0.91 | 0.007 (-0.007, 0.022) | 0.32 |
| CVR in white matter hyperintensity | -0.017 (-0.043, 0.009) | 0.196 | 0.004 (-0.023, 0.03) | 0.788 | -0.002 (-0.031, 0.028) | 0.916 | 0.003 (-0.037, 0.042) | 0.896 |
| CVR in subcortical gray matter | -0.018 (-0.048, 0.012) | 0.23 | 0.008 (-0.023, 0.039) | 0.613 | -0.006 (-0.041, 0.029) | 0.75 | 0.016 (-0.03, 0.063) | 0.488 |
| **Permeability surface area product, $10^{-4}min^{-1}$** | | | | | | | | |
| PS in normal appearing white matter | 0.022 (-0.527, 0.57) | 0.938 | 0.118 (-0.485, 0.72) | 0.7 | 0.092 (-0.537, 0.721) | 0.774 | -0.269 (-1.17, 0.631) | 0.556 |
| PS in white matter hyperintensity | 0.036 (-0.855, 0.926) | 0.937 | 0.643 (-0.331, 1.618) | 0.194 | 0.119 (-0.902, 1.141) | 0.818 | -1.152 (-2.605, 0.302) | 0.12 |
| PS in subcortical gray matter | -0.391 (-1.083, 0.302) | 0.267 | 0.696 (-0.061, 1.453) | 0.071 | -0.016 (-0.813, 0.782) | 0.969 | -0.412 (-1.552, 0.729) | 0.477 |
| **Plasma volume, $10^{-2}$** | | | | | | | | |
| $v_p$ in normal appearing white matter | 0.038 (-0.063, 0.138) | 0.461 | 0.025 (-0.085, 0.136) | 0.654 | 0.099 (-0.016, 0.214) | 0.09 | -0.062 (-0.227, 0.103) | 0.462 |
| $v_p$ in white matter hyperintensity | 0.124 (-0.079, 0.328) | 0.23 | 0.124 (-0.1, 0.348) | 0.277 | 0.11 (-0.124, 0.344) | 0.355 | 0.015 (-0.321, 0.35) | 0.932 |
| $v_p$ in subcortical gray matter | 0.01 (-0.182, 0.201) | 0.92 | 0.025 (-0.185, 0.236) | 0.812 | 0.172 (-0.046, 0.39) | 0.121 | -0.059 (-0.373, 0.256) | 0.714 |
| **Phase contrast MRI** | | | | | | | | |
| Internal carotid artery pulsatility index | -0.174 (-0.341, -0.007) | 0.041 | 0.08 (-0.098, 0.258) | 0.378 | 0.141 (-0.058, 0.339) | 0.164 | -0.27 (-0.639, 0.098) | 0.15 |
| Superior sagittal sinus pulsatility index | 0.033 (-0.064, 0.13) | 0.502 | -0.079 (-0.184, 0.026) | 0.139 | 0.165 (0.05, 0.281) | 0.005 | -0.103 (-0.292, 0.086) | 0.285 |
| CSF stroke volume, ml | -0.141 (-0.298, 0.016) | 0.078 | 0.013 (-0.154, 0.181) | 0.874 | -0.04 (-0.227, 0.147) | 0.673 | -0.176 (-0.475, 0.123) | 0.248 |

Abbreviations: ICA, internal carotid artery; MCA, middle cerebral artery; VA, vertebral artery; BA, basilar artery; CVR, cerebrovascular reactivity; PS, permeability surface area product; $v_p$, plasma volume; CSF, cerebrospinal fluid.

#### 2.4 Supplementary Table 4. Model comparison of linear vs. restricted cubic spline regressions across exposure–outcome pairs[a]

| Exposure | Outcome | Linear AIC | Linear BIC | Linear $R^2$ | RCS AIC | RCS BIC | RCS $R^2$ | Best Model | AIC Diff | BIC Diff | $R^2$ Diff |
|---|---|---|---|---|---|---|---|---|---|---|---|
| BA diameter | CVR in NAWM | -973.9 | -941.8 | 0.056 | -975.9 | -940.6 | 0.076 | RCS | -2.02 | 1.19 | 0.021 |
| VA diameter | CVR in NAWM | -975.1 | -943.1 | 0.062 | -975.6 | -940.4 | 0.075 | Linear | -0.53 | 2.67 | 0.013 |
| ICA diameter | CVR in NAWM | -974.2 | -942.2 | 0.057 | -973.2 | -938.0 | 0.063 | Linear | 1.01 | 4.21 | 0.005 |
| MCA diameter | CVR in NAWM | -976.2 | -944.1 | 0.067 | -974.3 | -939.1 | 0.068 | Linear | 1.87 | 5.07 | 0.001 |
| Mean arterial diameter | CVR in NAWM | -979.7 | -947.6 | 0.085 | -979.2 | -943.9 | 0.093 | Linear | 0.46 | 3.67 | 0.008 |
| BA diameter | CVR in WMH | -593.9 | -562.3 | 0.045 | -593.5 | -558.7 | 0.053 | Linear | 0.40 | 3.56 | 0.009 |
| VA diameter | CVR in WMH | -592.2 | -560.6 | 0.035 | -593.7 | -558.9 | 0.054 | Linear | -1.46 | 1.71 | 0.019 |
| ICA diameter | CVR in WMH | -590.1 | -558.5 | 0.024 | -589.1 | -554.3 | 0.029 | Linear | 1.07 | 4.24 | 0.005 |
| MCA diameter | CVR in WMH | -596.0 | -564.4 | 0.056 | -594.4 | -559.5 | 0.058 | Linear | 1.65 | 4.82 | 0.002 |
| Mean arterial diameter | CVR in WMH | -595.1 | -563.5 | 0.051 | -593.2 | -558.3 | 0.052 | Linear | 1.96 | 5.12 | 0.000 |
| BA diameter | CVR in SGM | -552.8 | -520.7 | 0.083 | -555.3 | -520.1 | 0.106 | RCS | -2.55 | 0.65 | 0.023 |
| VA diameter | CVR in SGM | -553.0 | -520.9 | 0.084 | -553.7 | -518.5 | 0.098 | Linear | -0.74 | 2.47 | 0.014 |
| ICA diameter | CVR in SGM | -552.9 | -520.9 | 0.084 | -551.1 | -515.9 | 0.085 | Linear | 1.81 | 5.02 | 0.001 |
| MCA diameter | CVR in SGM | -556.8 | -524.8 | 0.103 | -555.5 | -520.2 | 0.107 | Linear | 1.31 | 4.52 | 0.003 |
| Mean arterial diameter | CVR in SGM | -554.7 | -522.7 | 0.093 | -553.1 | -517.8 | 0.095 | Linear | 1.65 | 4.86 | 0.002 |
| BA diameter | PS in NAWM | 551.1 | 583.6 | 0.037 | 548.2 | 583.9 | 0.062 | RCS | -2.89 | 0.36 | 0.024 |
| VA diameter | PS in NAWM | 551.0 | 583.4 | 0.038 | 551.9 | 587.6 | 0.044 | Linear | 0.92 | 4.16 | 0.005 |
| ICA diameter | PS in NAWM | 551.3 | 583.8 | 0.036 | 553.0 | 588.7 | 0.038 | Linear | 1.67 | 4.91 | 0.002 |
| MCA diameter | PS in NAWM | 551.5 | 584.0 | 0.035 | 539.8 | 575.5 | 0.102 | RCS | -11.72 | -8.48 | 0.067 |
| Mean arterial diameter | PS in NAWM | 551.3 | 583.8 | 0.036 | 539.4 | 575.1 | 0.104 | RCS | -11.92 | -8.67 | 0.068 |
| BA diameter | PS in WMH | 735.7 | 768.2 | 0.027 | 737.2 | 772.9 | 0.029 | Linear | 1.47 | 4.72 | 0.003 |
| VA diameter | PS in WMH | 734.7 | 767.1 | 0.032 | 735.8 | 771.5 | 0.037 | Linear | 1.12 | 4.37 | 0.004 |
| ICA diameter | PS in WMH | 735.7 | 768.1 | 0.027 | 737.2 | 772.9 | 0.030 | Linear | 1.51 | 4.75 | 0.003 |

| | | | | | | | | | | | |
|---|---|---|---|---|---|---|---|---|---|---|---|
| MCA diameter | PS in WMH | 735.0 | 767.5 | 0.030 | 732.9 | 768.6 | 0.051 | RCS | -2.10 | 1.15 | 0.021 |
| Mean arterial diameter | PS in WMH | 735.7 | 768.2 | 0.027 | 732.3 | 768.0 | 0.054 | RCS | -3.40 | -0.15 | 0.027 |
| BA diameter | PS in SGM | 640.8 | 673.3 | 0.045 | 638.6 | 674.3 | 0.066 | RCS | -2.22 | 1.03 | 0.021 |
| VA diameter | PS in SGM | 640.9 | 673.4 | 0.044 | 640.6 | 676.3 | 0.056 | Linear | -0.37 | 2.88 | 0.012 |
| ICA diameter | PS in SGM | 639.4 | 671.9 | 0.052 | 641.4 | 677.1 | 0.052 | Linear | 1.95 | 5.19 | 0.000 |
| MCA diameter | PS in SGM | 641.2 | 673.7 | 0.043 | 639.0 | 674.7 | 0.064 | RCS | -2.24 | 1.01 | 0.021 |
| Mean arterial diameter | PS in SGM | 641.2 | 673.6 | 0.043 | 639.4 | 675.1 | 0.062 | Linear | -1.78 | 1.47 | 0.019 |
| BA diameter | $v_p$ in NAWM | -93.0 | -60.5 | 0.075 | -92.4 | -56.6 | 0.082 | Linear | 0.63 | 3.88 | 0.007 |
| VA diameter | $v_p$ in NAWM | -93.6 | -61.1 | 0.078 | -95.0 | -59.2 | 0.094 | Linear | -1.41 | 1.84 | 0.016 |
| ICA diameter | $v_p$ in NAWM | -93.6 | -61.1 | 0.078 | -91.7 | -56.0 | 0.079 | Linear | 1.90 | 5.15 | 0.000 |
| MCA diameter | $v_p$ in NAWM | -92.8 | -60.3 | 0.074 | -92.3 | -56.6 | 0.082 | Linear | 0.44 | 3.68 | 0.008 |
| Mean arterial diameter | $v_p$ in NAWM | -93.1 | -60.7 | 0.076 | -93.1 | -57.3 | 0.085 | Linear | 0.08 | 3.33 | 0.009 |
| BA diameter | $v_p$ in WMH | 174.5 | 207.0 | 0.079 | 176.3 | 212.0 | 0.080 | Linear | 1.81 | 5.06 | 0.001 |
| VA diameter | $v_p$ in WMH | 176.4 | 208.9 | 0.069 | 178.3 | 214.0 | 0.070 | Linear | 1.86 | 5.11 | 0.001 |
| ICA diameter | $v_p$ in WMH | 176.4 | 208.9 | 0.069 | 177.6 | 213.3 | 0.073 | Linear | 1.20 | 4.45 | 0.004 |
| MCA diameter | $v_p$ in WMH | 175.5 | 207.9 | 0.074 | 176.8 | 212.5 | 0.077 | Linear | 1.37 | 4.62 | 0.003 |
| Mean arterial diameter | $v_p$ in WMH | 175.6 | 208.1 | 0.073 | 177.6 | 213.3 | 0.074 | Linear | 1.94 | 5.19 | 0.000 |
| BA diameter | $v_p$ in SGM | 151.6 | 184.1 | 0.080 | 153.1 | 188.8 | 0.083 | Linear | 1.41 | 4.65 | 0.003 |
| VA diameter | $v_p$ in SGM | 151.0 | 183.5 | 0.083 | 152.2 | 187.9 | 0.087 | Linear | 1.13 | 4.37 | 0.004 |
| ICA diameter | $v_p$ in SGM | 149.5 | 181.9 | 0.090 | 151.4 | 187.1 | 0.091 | Linear | 1.89 | 5.14 | 0.001 |
| MCA diameter | $v_p$ in SGM | 150.3 | 182.7 | 0.087 | 150.5 | 186.2 | 0.095 | Linear | 0.24 | 3.48 | 0.008 |
| Mean arterial diameter | $v_p$ in SGM | 151.5 | 184.0 | 0.081 | 152.9 | 188.6 | 0.084 | Linear | 1.37 | 4.62 | 0.003 |
| BA diameter | Arterial PI | 120.3 | 153.2 | 0.233 | 118.6 | 154.7 | 0.247 | Linear | -1.73 | 1.56 | 0.014 |
| VA diameter | Arterial PI | 121.1 | 153.9 | 0.230 | 122.6 | 158.7 | 0.232 | Linear | 1.54 | 4.82 | 0.002 |
| ICA diameter | Arterial PI | 119.7 | 152.6 | 0.235 | 121.0 | 157.1 | 0.238 | Linear | 1.29 | 4.57 | 0.003 |

| | | | | | | | | | | | |
|---|---|---|---|---|---|---|---|---|---|---|---|
| MCA diameter | Arterial PI | 120.0 | 152.8 | 0.234 | 121.9 | 158.1 | 0.234 | Linear | 1.95 | 5.23 | 0.000 |
| Mean arterial diameter | Arterial PI | 121.1 | 153.9 | 0.230 | 121.8 | 157.9 | 0.235 | Linear | 0.73 | 4.02 | 0.005 |
| BA diameter | Venous PI | -88.8 | -55.7 | 0.261 | -91.4 | -55.0 | 0.278 | RCS | -2.60 | 0.71 | 0.017 |
| VA diameter | Venous PI | -88.6 | -55.5 | 0.260 | -91.8 | -55.4 | 0.279 | RCS | -3.23 | 0.08 | 0.019 |
| ICA diameter | Venous PI | -92.3 | -59.3 | 0.274 | -90.4 | -54.0 | 0.274 | Linear | 1.96 | 5.26 | 0.000 |
| MCA diameter | Venous PI | -89.7 | -56.7 | 0.265 | -88.3 | -51.9 | 0.267 | Linear | 1.43 | 4.74 | 0.002 |
| Mean arterial diameter | Venous PI | -88.7 | -55.6 | 0.261 | -88.0 | -51.6 | 0.265 | Linear | 0.69 | 4.00 | 0.005 |
| BA diameter | CSF stroke volume | 95.1 | 128.0 | 0.022 | 97.1 | 133.3 | 0.022 | Linear | 1.97 | 5.26 | 0.000 |
| VA diameter | CSF stroke volume | 95.7 | 128.6 | 0.019 | 97.4 | 133.6 | 0.021 | Linear | 1.71 | 5.00 | 0.001 |
| ICA diameter | CSF stroke volume | 93.0 | 125.9 | 0.033 | 95.0 | 131.2 | 0.033 | Linear | 2.00 | 5.29 | 0.000 |
| MCA diameter | CSF stroke volume | 90.7 | 123.6 | 0.044 | 92.6 | 128.8 | 0.044 | Linear | 1.97 | 5.26 | 0.000 |
| Mean arterial diameter | CSF stroke volume | 92.8 | 125.7 | 0.034 | 93.7 | 129.8 | 0.039 | Linear | 0.88 | 4.17 | 0.005 |

[a] Each exposure-outcome pair was fitted with (i) a linear regression and (ii) an RCS regression, both adjusted for age, sex, hypertension, diabetes mellitus, hyperlipidemia, smoking, and mean blood pressure. Linear AIC/BIC/$R^2$: model fit indices from adjusted linear regressions. RCS AIC/BIC/$R^2$: fit indices from restricted cubic spline regressions. AIC Diff, BIC Diff: RCS - Linear values; negative indicates RCS fit better. $R^2$ Diff: RCS $R^2$ - Linear $R^2$. Best Model: chosen based on AIC. If AIC difference >2, the model with lower AIC was selected; if |AIC difference| $\leqslant 2$, RCS was selected when the non-linearity test was significant ($p<0.05$), otherwise Linear was retained. Abbreviations: NAWM, normal-appearing white matter; WMH, white matter hyperintensity; SGM, subcortical gray matter; CVR, cerebrovascular reactivity; PS, permeability surface area product; $v_p$, plasma volume; PI, pulsatility index; CSF, cerebrospinal fluid.

#### 2.5 Supplementary Table 5. Model fit statistics comparing models with and without the region×diameter interaction[a]

| | CVR | PS |
|---|---|---|
| AIC (with interaction) | -1966.3 | 1860.96 |
| AIC (without interaction) | -1967.58 | 1854.08 |
| BIC (with interaction) | -1901.96 | 1939.18 |
| BIC (without interaction) | -1911.81 | 1914.92 |
| LogLik (with interaction | 998.15 | -912.48 |
| LogLik (without interaction) | 996.79 | -913.04 |
| $\chi^2$ (LRT) | 2.728 | 1.123 |
| df | 2 | 4 |
| p-value | 0.256 | 0.891 |

[a] Models for CVR were fitted using linear mixed-effects models, while models for PS were fitted using restricted cubic spline mixed-effects models. Both included random intercepts for participant ID to account for within-subject correlation and were adjusted for age, sex, hypertension, diabetes mellitus, hyperlipidemia, smoking, and mean blood pressure. AIC, BIC: Akaike and Bayesian Information Criteria; lower values indicate better model fit. LogLik: model log-likelihood. $\chi^2$ (LRT), df, and p-value: results from likelihood ratio tests comparing the full model (with interaction) against the reduced model (without interaction). Abbreviations: CVR, cerebrovascular reactivity; PS, permeability surface area product; df, degrees of freedom; LRT, likelihood ratio test.

**2.6 Supplementary Figure 3. Associations between mean arterial diameter and cerebrovascular function across regions.** (A) Association of mean arterial diameter (z-score) with cerebrovascular reactivity (CVR) in normal-appearing white matter (NAWM, blue), white matter hyperintensities (WMH, orange), and subcortical gray matter (SGM, green), estimated using linear mixed-effects models. (B) Association of mean arterial diameter (z-score) with permeability surface area product (PS) in NAWM, WMH, and SGM, estimated using restricted cubic spline mixed-effects models. Both models included random intercepts for participant ID and were adjusted for age, sex, hypertension, diabetes mellitus, hyperlipidemia, smoking, and mean blood pressure. Abbreviations: NAWM, normal-appearing white matter; WMH, white matter hyperintensity; SGM, subcortical gray matter.

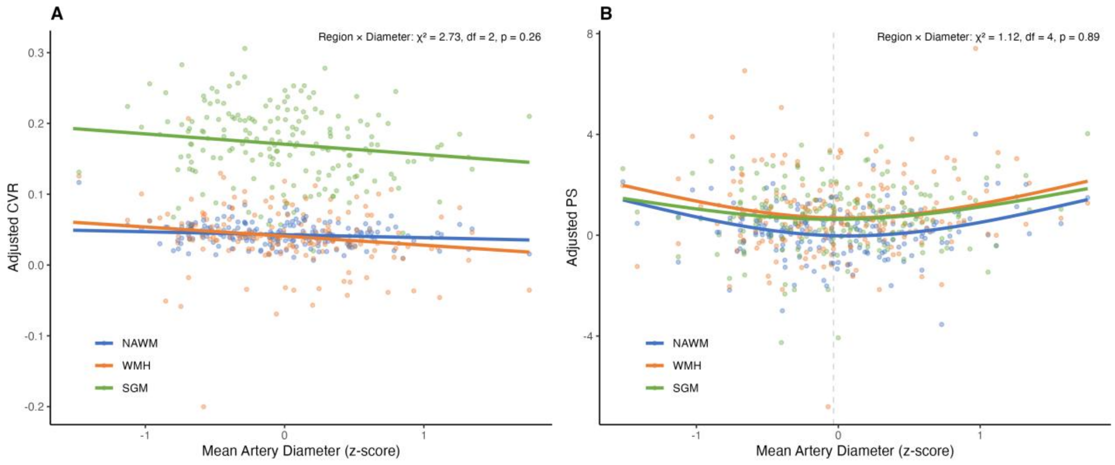

#### 2.7 Supplementary Table 6. Linear regressions of intracranial arterial diameters with plasma volume[a]

| | $v_p$ in NAWM | | $v_p$ in WMH | | $v_p$ in SGM | |
|---|---|---|---|---|---|---|
| | β (95% CI) | P value | β (95% CI) | P value | β (95% CI) | P value |
| Mean diameter | 0.02 (-0.033, 0.072) | 0.464 | -0.048 (-0.154, 0.059) | 0.376 | 0.020 (-0.08, 0.12) | 0.692 |
| ICA diameter | 0.022 (-0.021, 0.064) | 0.319 | -0.010 (-0.097, 0.076) | 0.815 | 0.059 (-0.022, 0.14) | 0.15 |
| MCA diameter | -0.018 (-0.101, 0.064) | 0.661 | -0.082 (-0.25, 0.086) | 0.335 | -0.092 (-0.249, 0.065) | 0.249 |
| BA diameter | 0.0130 (-0.028, 0.054) | 0.527 | -0.056 (-0.138, 0.026) | 0.176 | 0.002 (-0.076, 0.079) | 0.966 |
| VA diameter | 0.021 (-0.022, 0.065) | 0.335 | 0.009 (-0.079, 0.098) | 0.832 | 0.032 (-0.05, 0.115) | 0.441 |

[a] Models were adjusted for age, sex, hypertension, diabetes mellitus, hyperlipidemia, smoking, and mean blood pressure. Mean arterial diameter was standardized (z-score), so β values represent change in outcome per 1-SD larger mean diameter; vessel-specific diameters were analyzed in mm. Abbreviations: CI, confidential interval; NAWM, normal-appearing white matter; WMH, white matter hyperintensity; SGM, subcortical gray matter; ICA, internal carotid artery; MCA, middle cerebral artery; BA, basilar artery; VA, vertebral artery; $v_p$, plasma volume.

#### 2.8 Supplementary Table 7. Linear regressions of intracranial arterial diameters with pulsatility[a]

| | Internal carotid artery pulsatility index | | Superior sagittal sinus pulsatility index | | CSF stroke volume at the foramen magnum | |
|---|---|---|---|---|---|---|
| | β (95% CI) | P value | β (95% CI) | P value | β (95% CI) | P value |
| Mean diameter | -0.008 (-0.100, 0.084) | 0.869 | 0.010 (-0.043, 0.064) | 0.709 | 0.072 (-0.013, 0.157) | 0.097 |
| ICA diameter | 0.042 (-0.030, 0.113) | 0.249 | 0.040 (-0.001, 0.082) | 0.057 | 0.054 (-0.013, 0.120) | 0.111 |
| MCA diameter | -0.078 (-0.226, 0.070) | 0.301 | -0.047 (-0.132, 0.039) | 0.282 | 0.153 (0.016, 0.290) | 0.029 |
| BA diameter | -0.030 (-0.099, 0.039) | 0.385 | -0.011 (-0.051, 0.029) | 0.589 | -0.024 (-0.088, 0.040) | 0.460 |
| VA diameter | -0.007 (-0.078, 0.065) | 0.855 | 0.004 (-0.038, 0.045) | 0.864 | 0.002 (-0.064, 0.068) | 0.949 |

[a] Models were adjusted for age, sex, hypertension, diabetes mellitus, hyperlipidemia, smoking, and mean blood pressure. Mean arterial diameter was standardized (z-score), so β values represent change in outcome per 1-SD larger mean diameter; vessel-specific diameters were analyzed in mm. Abbreviations: CI, confidential interval; NAWM, normal-appearing white matter; WMH, white matter hyperintensity; SGM, subcortical gray matter; ICA, internal carotid artery; MCA, middle cerebral artery; BA, basilar artery; VA, vertebral artery.

#### 2.9 Supplementary Table 8 Mediation analyses of the associations between basilar artery dolichoectasia (exposure) and cerebral small vessel disease markers (outcome), with cerebrovascular reactivity in normal-appearing white matter as the mediator

| | Indirect effect | | Direct effect | | Total Effect | | Proportion Mediated | |
|---|---|---|---|---|---|---|---|---|
| | β (95% CI) | P value | β (95% CI) | P value | β (95% CI) | P value | β (95% CI) | P value |
| Baseline summary SVD score | 0.120 (0.010, 0.286) | 0.024 | 0.440 (-0.125, 0.977) | 0.126 | 0.560 (-0.009, 1.094) | 0.054 | 0.214 (-0.134, 1.221) | 0.077 |
| Baseline number of lacunes | 0.279 (0.026, 0.646) | 0.026 | 1.758 (0.285, 3.379) | 0.019 | 2.037 (0.473, 3.768) | 0.007 | 0.137 (0.014, 0.460) | 0.032 |
| Baseline number of microbleeds | 0.648 (0.082, 1.644) | 0.006 | 2.644 (-0.552, 6.787) | 0.196 | 3.292 (-0.306, 8.169) | 0.128 | 0.197 (-1.091, 1.268) | 0.128 |
| Baseline WMH volume | 0.033 (-0.003, 0.086) | 0.087 | 0.203 (0.056, 0.353) | 0.005 | 0.236 (0.081, 0.397) | 0.003 | 0.141 (-0.020, 0.416) | 0.088 |
| Baseline PVS volume | -0.007 (-0.032, 0.016) | 0.458 | 0.110 (0.016, 0.205) | 0.024 | 0.102 (0.010, 0.193) | 0.028 | -0.073 (-0.650, 0.260) | 0.472 |
| NAWM→WMH at 1 year | 0.053 (0.009, 0.116) | 0.007 | 0.215 (0.044, 0.384) | 0.016 | 0.268 (0.088, 0.447) | 0.004 | 0.199 (0.039, 0.559) | 0.011 |
| PVS volume growth at 1 year | 0.26 (0.051, 0.564) | 0.006 | 0.609 (-0.321, 1.456) | 0.176 | 0.869 (-0.093, 1.816) | 0.078 | 0.299 (-0.643, 1.625) | 0.081 |
| Incident infarct in 1 year | 0.008 (-0.034, 0.050) | 0.725 | 0.194 (-0.009, 0.393) | 0.059 | 0.202 (-0.002, 0.399) | 0.052 | 0.041 (-0.289, 0.416) | 0.725 |

*Models were adjusted for age, sex, hypertension, diabetes mellitus, hyperlipidemia, smoking, and mean blood pressure. Abbreviations: CI, confidential interval; NAWM, normal appearing white matter; WMH, white matter hyperintensity; PVS, perivascular space.

#### 2.10 Supplementary Table 9. Sensitivity analysis using maximum instead of mean diameters in relation to cerebrovascular function[a]

| | CVR in NAWM | | CVR in WMH | | CVR in SGM | |
|---|---|---|---|---|---|---|
| | β (95% CI) | P value | β (95% CI) | P value | β (95% CI) | P value |
| max ICA diameter | -0.0015 (-0.0054, 0.0025) | 0.465 | 0.0011 (-0.0095, 0.0117) | 0.834 | -0.0027 (-0.0152, 0.0097) | 0.664 |
| max MCA diameter | -0.0094 (-0.0166, -0.0022) | 0.011 | -0.0208 (-0.0404, -0.0012) | 0.038 | -0.0253 (-0.0482, -0.0023) | 0.031 |
| max VA diameter | -0.0024 (-0.0056, 0.0007) | 0.122 | -0.0062 (-0.0148, 0.0024) | 0.159 | -0.0008 (-0.0107, 0.0091) | 0.876 |
| | PS in NAWM | | PS in WMH | | PS in SGM | |
| | β (95% CI) | P value | β (95% CI) | P value | β (95% CI) | P value |
| max ICA diameter | 0.0618 (-0.1767, 0.3004) | 0.979 | -0.0625 (-0.4499, 0.3250) | 0.862 | 0.2143 (-0.0864, 0.5151) | 0.551 |
| max MCA diameter | 0.1492 (-0.2172, 0.5157) | <0.001 | -0.1445 (-0.7402, 0.4512) | 0.016 | -0.0520 (-0.5170, 0.4130) | 0.009 |
| max VA diameter | -0.0499 (-0.2465, 0.1467) | 0.220 | 0.0879 (-0.2313, 0.4071) | 0.469 | -0.0496 (-0.2988, 0.1995) | 0.270 |
| | $v_p$ in NAWM | | $v_p$ in WMH | | $v_p$ in SGM | |
| | β (95% CI) | P value | β (95% CI) | P value | β (95% CI) | P value |
| max ICA diameter | 0.0212 (-0.0225, 0.0649) | 0.340 | -0.0104 (-0.0994, 0.0785) | 0.818 | 0.0562 (-0.0268, 0.1391) | 0.183 |
| max MCA diameter | -0.0025 (-0.0699, 0.0649) | 0.942 | -0.0417 (-0.1784, 0.0950) | 0.548 | -0.0550 (-0.1829, 0.0729) | 0.397 |
| max VA diameter | 0.0290 (-0.0069, 0.0648) | 0.113 | 0.0270 (-0.0462, 0.1002) | 0.468 | 0.0202 (-0.0484, 0.0889) | 0.562 |
| | Internal carotid artery pulsatility index | | Superior sagittal sinus pulsatility index | | CSF stroke volume at the foramen magnum | |
| | β (95% CI) | P value | β (95% CI) | P value | β (95% CI) | P value |
| max ICA diameter | 0.0504 (-0.0227, 0.1236) | 0.176 | 0.0409 (-0.0013, 0.0832) | 0.058 | 0.0477 (-0.0201, 0.1155) | 0.167 |
| max MCA diameter | -0.0985 (-0.2230, 0.0260) | 0.120 | -0.0790 (-0.1512, -0.0067) | 0.032 | 0.1117 (-0.0046, 0.2279) | 0.060 |
| max VA diameter | -0.0226 (-0.0822, 0.0369) | 0.454 | -0.0110 (-0.0458, 0.0237) | 0.531 | -0.0033 (-0.0584, 0.0519) | 0.907 |

[a]Models were adjusted for age, sex, hypertension, diabetes mellitus, hyperlipidemia, smoking, and mean blood pressure. Abbreviations: CI, confidential interval; NAWM, normal-appearing white matter; WMH, white matter hyperintensity; SGM, subcortical gray matter; ICA, internal carotid artery; MCA, middle cerebral artery; VA, vertebral artery; CVR, cerebrovascular reactivity; PS, permeability surface area product; vP, plasma volume; CSF, cerebrospinal fluid.

#### 2.11 Supplementary Table 10. Sensitivity analysis additionally adjusting for large artery stenosis in the associations of intracranial arterial diameters with cerebrovascular function[a]

| | CVR in NAWM | | CVR in WMH | | CVR in SGM | |
|---|---|---|---|---|---|---|
| | β (95% CI) | P value | β (95% CI) | P value | β (95% CI) | P value |
| Mean arterial diameter | -0.0067 (-0.0115, -0.0019) | 0.006 | -0.0151 (-0.0282, -0.0020) | 0.024 | -0.0112 (-0.0266, 0.0043) | 0.156 |
| ICA diameter | -0.0031 (-0.0070, 0.0009) | 0.127 | -0.0015 (-0.0122, 0.0091) | 0.775 | -0.0034 (-0.0159, 0.0091) | 0.592 |
| MCA diameter | -0.0087 (-0.0169, -0.0004) | 0.041 | -0.0274 (-0.0497, -0.0052) | 0.016 | -0.0267 (-0.0530, -0.0005) | 0.046 |
| BA diameter | -0.0026 (-0.0062, 0.0010) | 0.156 | -0.0102 (-0.0202, -0.0002) | 0.046 | -0.0018 (-0.0133, 0.0097) | 0.756 |
| VA diameter | -0.0034 (-0.0070, 0.0003) | 0.072 | -0.0070 (-0.0170, 0.0030) | 0.169 | -0.0031 (-0.0147, 0.0086) | 0.601 |
| | PS in NAWM | | PS in WMH | | PS in SGM | |
| | β (95% CI) | P value | β (95% CI) | P value | β (95% CI) | P value |
| Mean arterial diameter | 0.0653 (-0.2215, 0.3522) | <0.001 | -0.0235 (-0.4895, 0.4426) | 0.024 | 0.0935 (-0.2698, 0.4569) | 0.059 |
| ICA diameter | 0.0586 (-0.1749, 0.2921) | 0.575 | -0.0400 (-0.4193, 0.3394) | 0.494 | 0.2048 (-0.0896, 0.4993) | 0.830 |
| MCA diameter | 0.0490 (-0.4034, 0.5015) | <0.001 | -0.3054 (-1.0387, 0.4279) | 0.050 | -0.1403 (-0.7130, 0.4325) | 0.046 |
| BA diameter | -0.0744 (-0.2961, 0.1474) | 0.032 | 0.0035 (-0.3570, 0.3639) | 0.482 | -0.1120 (-0.3927, 0.1688) | 0.046 |
| VA diameter | 0.0922 (-0.1460, 0.3304) | 0.313 | 0.1973 (-0.1890, 0.5836) | 0.363 | 0.1105 (-0.1913, 0.4123) | 0.136 |
| | $v_p$ in NAWM | | $v_p$ in WMH | | $v_p$ in SGM | |
| | β (95% CI) | P value | β (95% CI) | P value | β (95% CI) | P value |
| Mean arterial diameter | 0.0200 (-0.0325, 0.0724) | 0.454 | -0.0461 (-0.1516, 0.0594) | 0.389 | 0.0206 (-0.0795, 0.1206) | 0.685 |
| ICA diameter | 0.0234 (-0.0193, 0.0660) | 0.281 | -0.0035 (-0.0895, 0.0826) | 0.937 | 0.0613 (-0.0197, 0.1423) | 0.137 |
| MCA diameter | -0.0183 (-0.1011, 0.0645) | 0.663 | -0.0818 (-0.2480, 0.0845) | 0.333 | -0.0918 (-0.2491, 0.0654) | 0.251 |
| BA diameter | 0.0141 (-0.0265, 0.0547) | 0.495 | -0.0524 (-0.1338, 0.0290) | 0.206 | 0.0028 (-0.0746, 0.0802) | 0.943 |
| VA diameter | 0.0191 (-0.0244, 0.0627) | 0.387 | 0.0001 (-0.0877, 0.0880) | 0.998 | 0.0301 (-0.0530, 0.1132) | 0.476 |
| | Internal carotid artery pulsatility index | | Superior sagittal sinus pulsatility index | | CSF stroke volume at the foramen magnum | |
| | β (95% CI) | P value | β (95% CI) | P value | β (95% CI) | P value |

| | | | | | | |
|---|---|---|---|---|---|---|
| Mean arterial diameter | -0.010 (-0.102, 0.082) | 0.829 | 0.010 (-0.044, 0.064) | 0.708 | 0.069 (-0.017, 0.154) | 0.114 |
| ICA diameter | 0.039 (-0.033, 0.111) | 0.291 | 0.041 (-0.001, 0.083) | 0.055 | 0.049 (-0.017, 0.116) | 0.145 |
| MCA diameter | -0.080 (-0.228, 0.068) | 0.288 | -0.047 (-0.133, 0.039) | 0.283 | 0.150 (0.013, 0.287) | 0.032 |
| BA diameter | -0.031 (-0.100, 0.038) | 0.381 | -0.011 (-0.051, 0.029) | 0.591 | -0.026 (-0.090, 0.038) | 0.427 |
| VA diameter | -0.005 (-0.077, 0.067) | 0.892 | 0.004 (-0.038, 0.045) | 0.865 | 0.004 (-0.062, 0.070) | 0.896 |

[a] Models were adjusted for age, sex, hypertension, diabetes mellitus, hyperlipidemia, smoking, and mean blood pressure. Mean arterial diameter was standardized (z-score), so β values represent change in outcome per 1-SD larger mean diameter; vessel-specific diameters were analyzed in mm. Abbreviations: CI, confidential interval; NAWM, normal-appearing white matter; WMH, white matter hyperintensity; SGM, subcortical gray matter; ICA, internal carotid artery; MCA, middle cerebral artery; BA, basilar artery; VA, vertebral artery; CVR, cerebrovascular reactivity; PS, permeability surface area product; $v_p$, plasma volume; CSF, cerebrospinal fluid.
